# Pre-pandemic national immunisation programme strength and health workforce capacity improved routine immunisation resilience during the COVID-19 pandemic

**DOI:** 10.1101/2024.10.09.24315115

**Authors:** Beth Evans, Laurent Kaiser, Olivia Keiser, Thibaut Jombart

## Abstract

**Background:** The adverse impact of the COVID-19 pandemic on Routine Immunisation (RI) coverage has been well-documented: most countries experienced backsliding or stagnation in coverage. Qualitative surveys indicated potential causes of such declines, including reduced health care seeking behaviour, lockdowns, and overwhelmed health systems.

**Methods:** We investigate determinants of RI resilience during COVID-19 at a national level for 119 countries from 2020 to 2022, using publicly available data on pre-pandemic immunisation programme performance, health workforce capacity, health systems strength, health financing, global health security preparedness, COVID-19 burden, COVID-19 containment, economic, and health policy responses, population mobility changes, and country wealth. We employ a mixed methods approach: stepwise linear regression based on a causal inference framework, and Random Forest regression to identify potential nonlinear interactions and collinear effects.

**Results:** We provide evidence that stronger pre-pandemic immunisation programmes and more health workers, once above minimum thresholds (about 83% Diphtheria-Tetanus-Pertussis third-dose coverage and 60 health workers per 10,000 population), are associated with improved RI resilience. Random Forest analysis suggests health financing and health system strength impact RI resilience. COVID-19 vaccinations and pandemic policies were not associated with RI coverage changes, implying – reassuring – these acute responses did not interrupt routine services. In addition to these findings, a large fraction of variation in pandemic RI resilience remains unexplained, highlighting the need for further research on RI performance determinants.

**Conclusion:** Our findings underscore the role of robust immunisation programmes and sufficiently sized health workforces in mitigating RI disruption during global health crises once above minimum thresholds. Reassuringly, we do not find evidence that COVID-19 vaccination campaigns nor pandemic containment policies impacted RI performance – counter to qualitative survey indications. We encourage continued efforts to identify RI disruption determinants to inform the evidence base for public health practitioners globally.

---

1. **Introduction**

The COVID-19 pandemic, declared by the World Health Organization (WHO) from March 11^th^ 2020 to May 5^th^ 2023, caused widespread adverse effects on health, health systems, lifestyles, and economies worldwide. Many health systems were overwhelmed by COVID-19 cases and reported disrupted essential health service provision [1–3]. On the demand side, health care utilisation reduced by one-third during the pandemic, indicating changed health seeking behaviours [4]. Adoption of unprecedented non-pharmaceutical interventions (NPIs) such as lockdowns, school, workplace and border closures interrupted normal internal and international mobility [5]. These NPIs aimed to reduce COVID-19 transmission and impact, but potentially also influenced absolute or perceived access to health services.

Routine immunisation (RI) is an essential preventative intervention, referring to vaccinations given to infants under 24-months. RI schedules vary by country with 10 vaccines recommended by the WHO for all RI programmes and more recommended for certain regions or high-risk populations [6]. RI is estimated to prevent 3.5 to five million deaths per year [7]. Diphtheria-tetanus-pertussis containing vaccines third-dose (DTP3) coverage is used as a standardised tracker of immunisation programme performance over time and across countries [8]. Initial qualitative reporting [1–3] and subsequent quantitative modelling [9,10] described worldwide reductions in RI coverage during the pandemic. Using our previously published methodology [10], updated for the latest WHO/UNICEF Estimates of National Immunisation Coverage (WUENIC) estimates [11] these global DTP3 reductions were: 2.7% in 2020, 3.2% in 2021, and 2.5% in 2022. Most countries exhibited sustained declines or coverage stagnation, whilst some rebounded in later years, and only a minority achieved year-on-year coverage improvements [10]. Why were some immunisation programmes more resilient than others? Understanding the drivers could help prevent vaccine-preventable outbreaks and deaths during future epidemics or pandemics.

Significant research has been undertaken on determinants of immunisation system performance, within countries (e.g., [12]), and at a country-level (e.g., [12,13]). Previous research classified determinants along three dimensions: facility readiness – encompassing supply and workforce availability and quality, access to services e.g., distance to health facilities, and intent to vaccinate including demand-side norms such as vaccine hesitancy [14]. During the pandemic, qualitative studies explored pandemic RI disruption via questionnaires, interviews, and focus groups, with caregivers and health worker across single-countries e.g., Indonesia [15], and regions [16,17]. These surveys highlighted fear of contracting COVID-19, lack of public transport, and lockdowns as key drivers reducing health seeking behaviour during the pandemic. Health facility closures and insufficient health workers were cited as barriers to service delivery. RI disruptions due to COVID-19 vaccine rollout were reported in almost half of responding countries [3], and increased vaccine hesitancy was reported through parental surveys [18].

Our research focuses on the direct and indirect effects of the pandemic on RI, aiming to quantitatively evaluate insights from the aforementioned surveys, and identify country-level factors associated with changes in RI coverage during the COVID-19 pandemic to provide direction on maintaining robust immunisation systems in times of crises.

## 2. Material and methods

### 2.1 Conceptual framework

We build on a conceptual framework developed by Philips et al. (2017) – a literature review of 78 articles investigating coverage determinants in low- and middle-income countries [14], reproduced in Supplementary Materials section A, which we adapt to the pandemic context and data availability. See **Figure 1** for the resulting causal inference framework which maps identified available datasets to the conceptual framework. Details on each independent variable are summarised in **Table 1**.

**Figure 1.**
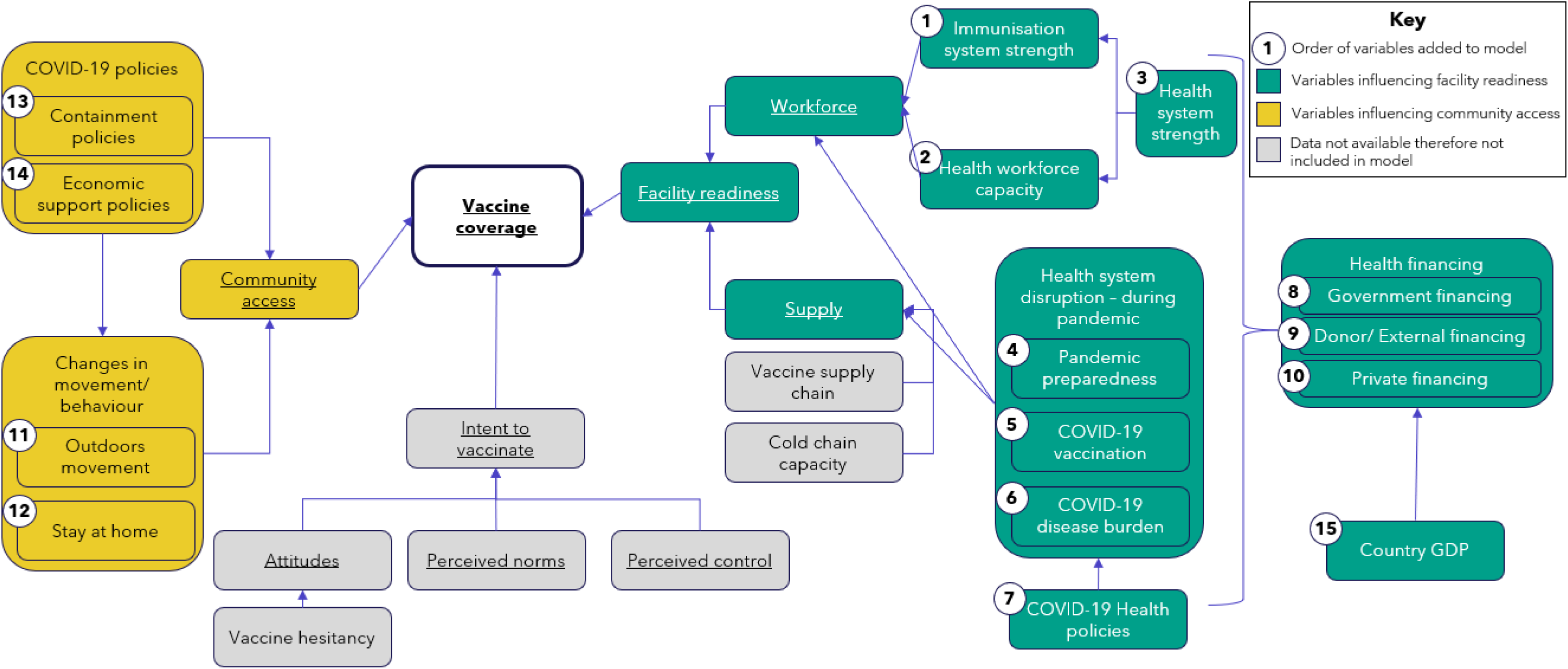
The causal inference framework for this research. This builds on research by Phillips et al. (2017), adapted for the pandemic context and available datasets. Numbers indicate proximity and order of investigating associations through linear modelling. Arrows indicate the directionality of relationship. Grey variables are not included in modelling due to lack of data. Underlining indicates variable included in source conceptual framework.

**Table 1.**
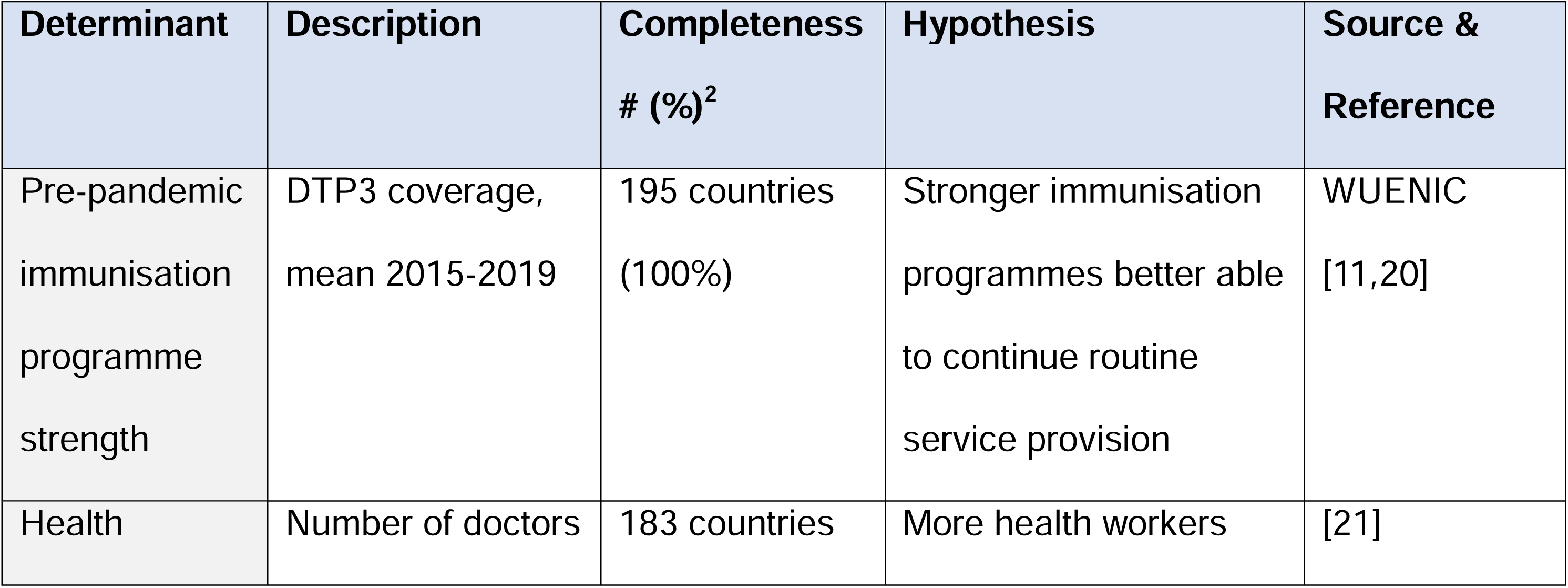
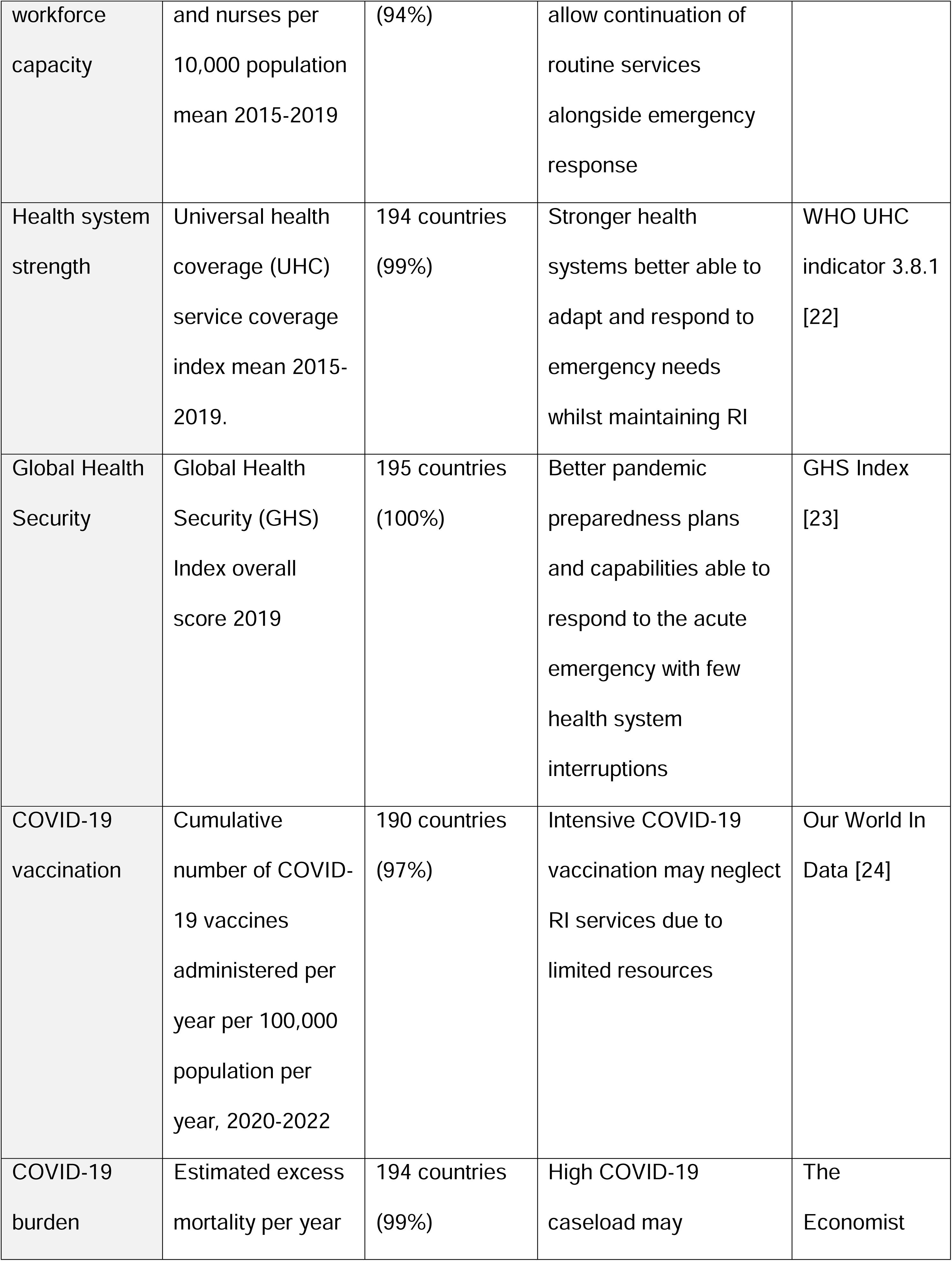
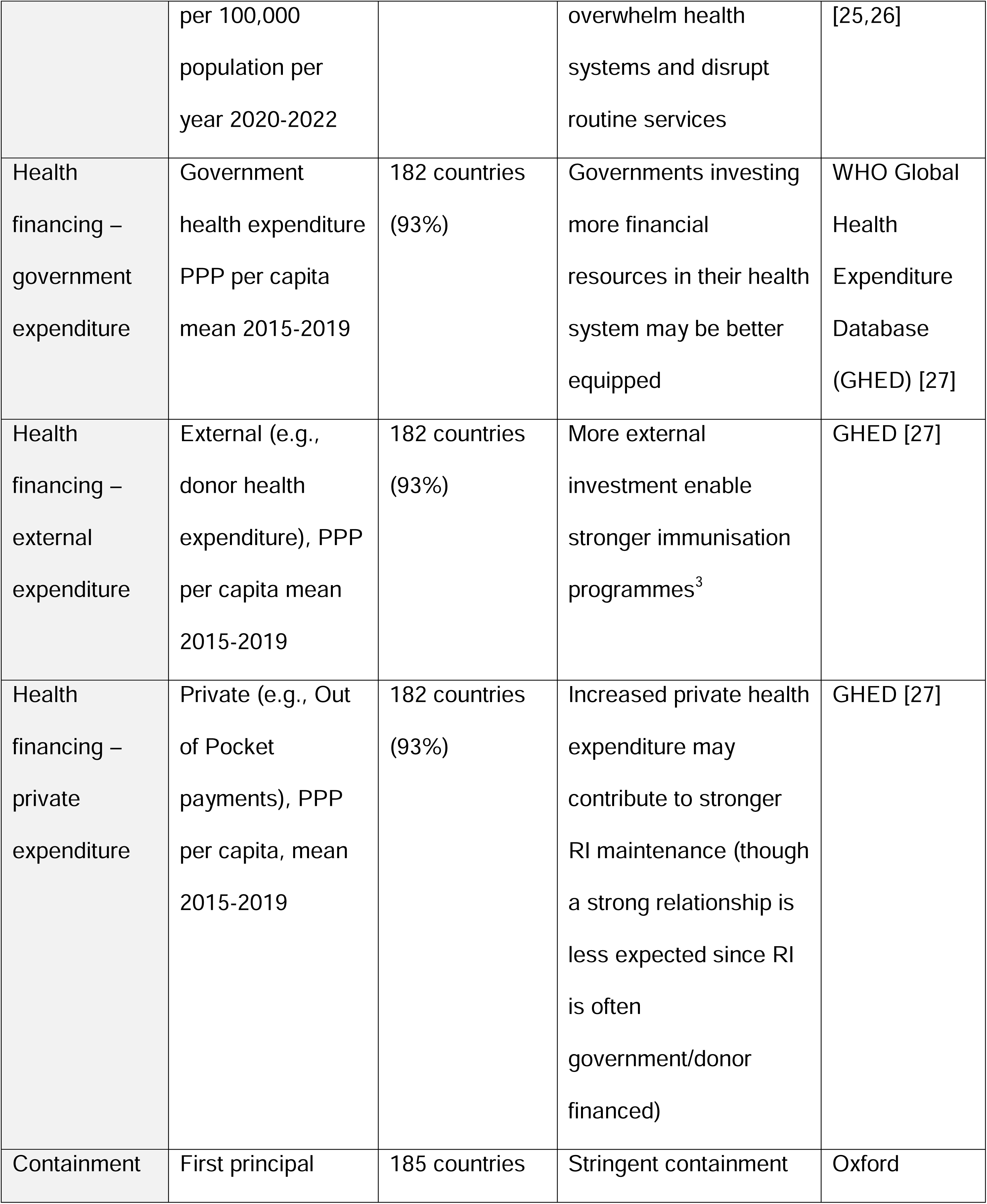
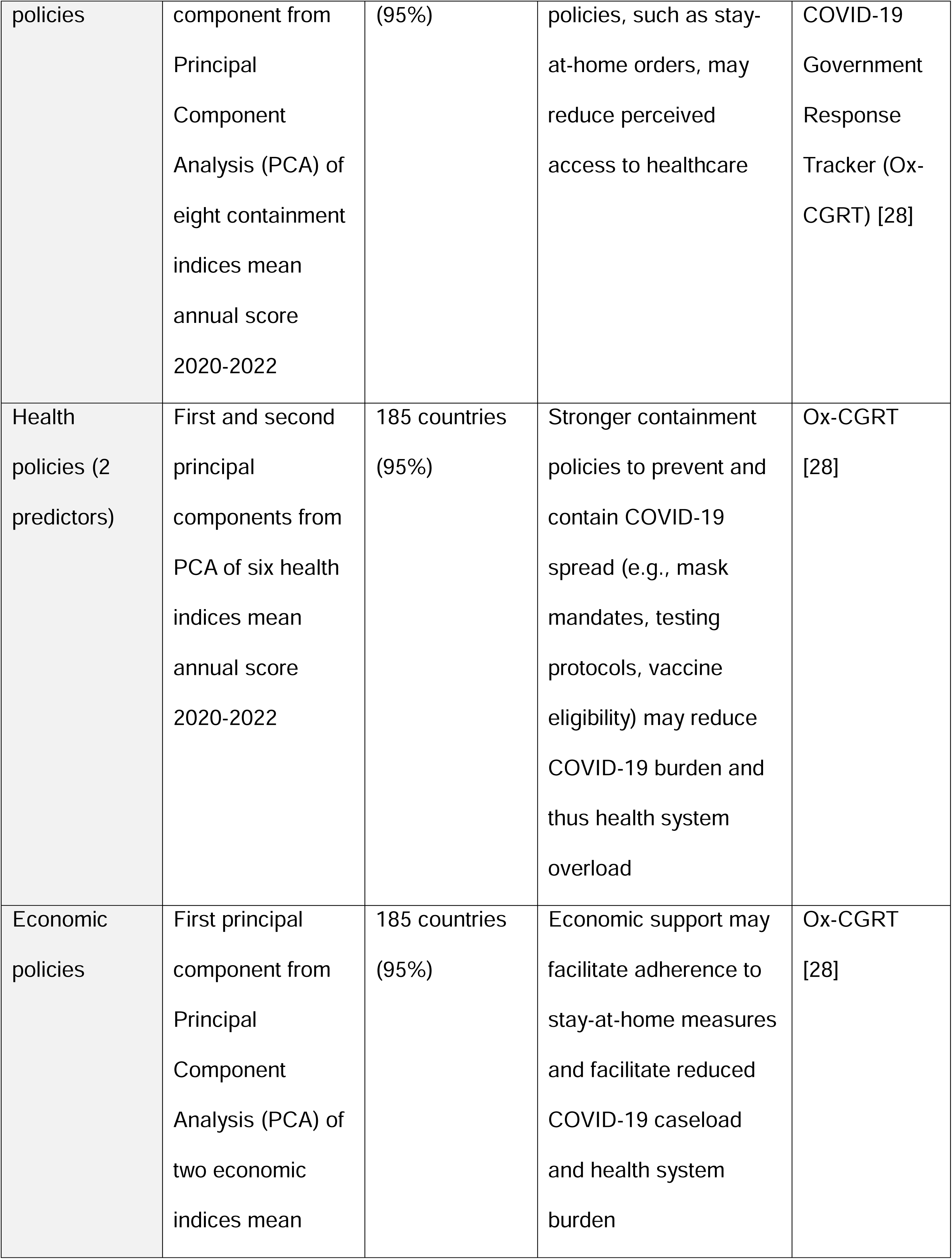
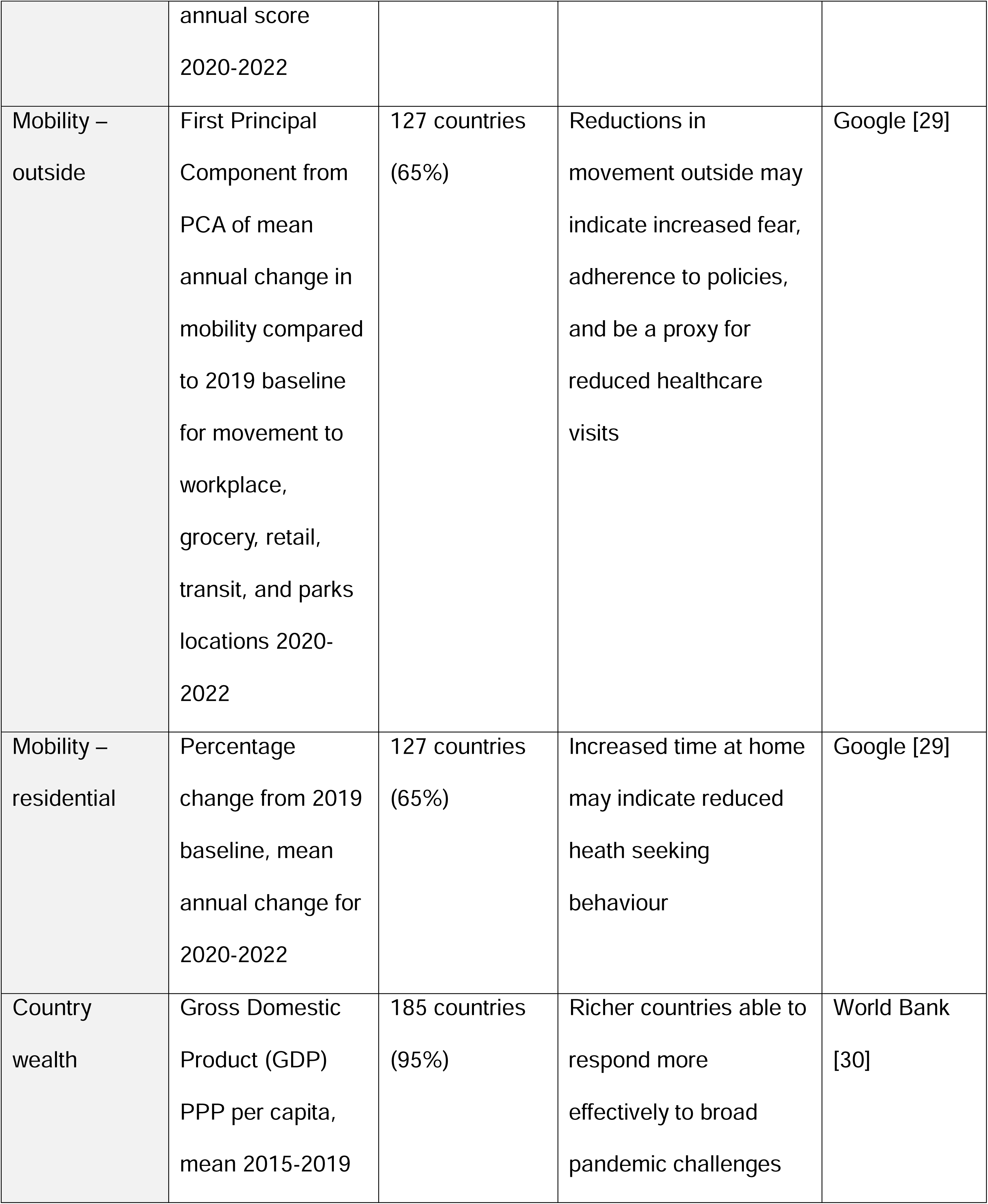
Summary of determinants, data source and description, completeness, and hypothesis for potential impact on routine immunisation services during the pandemic.

| Determinant | Description | Completeness<br># (%)<sup>2</sup> | Hypothesis | Source &<br>Reference |
| --- | --- | --- | --- | --- |
| Pre-pandemic<br>immunisation<br>programme<br>strength | DTP3 coverage,<br>mean 2015-2019 | 195 countries<br>(100%) | Stronger immunisation<br>programmes better able<br>to continue routine<br>service provision | WUENIC<br>[11,20] |
| Health | Number of doctors | 183 countries | More health workers | [21] |

<sup>1</sup> which converts different currencies and weights for costs of goods in a country to enable comparisons across countries
<sup>2</sup> Percentages are report in relation to the 195 countries included in WUENIC data

|  |  |  |  |  |
| --- | --- | --- | --- | --- |
| workforce capacity | and nurses per 10,000 population mean 2015-2019 | (94%) | allow continuation of routine services alongside emergency response |  |
| Health system strength | Universal health coverage (UHC) service coverage index mean 2015-2019. | 194 countries (99%) | Stronger health systems better able to adapt and respond to emergency needs whilst maintaining RI | WHO UHC indicator 3.8.1 [22] |
| Global Health Security | Global Health Security (GHS) Index overall score 2019 | 195 countries (100%) | Better pandemic preparedness plans and capabilities able to respond to the acute emergency with few health system interruptions | GHS Index [23] |
| COVID-19 vaccination | Cumulative number of COVID-19 vaccines administered per year per 100,000 population per year, 2020-2022 | 190 countries (97%) | Intensive COVID-19 vaccination may neglect RI services due to limited resources | Our World In Data [24] |
| COVID-19 burden | Estimated excess mortality per year | 194 countries (99%) | High COVID-19 caseload may | The Economist |
|  | per 100,000<br>population per<br>year 2020-2022 |  | overwhelm health<br>systems and disrupt<br>routine services | [25,26] |
| Health<br>financing –<br>government<br>expenditure | Government<br>health expenditure<br>PPP per capita<br>mean 2015-2019 | 182 countries<br>(93%) | Governments investing<br>more financial<br>resources in their health<br>system may be better<br>equipped | WHO Global<br>Health<br>Expenditure<br>Database<br>(GHED) [27] |
| Health<br>financing –<br>external<br>expenditure | External (e.g.,<br>donor health<br>expenditure), PPP<br>per capita mean<br>2015-2019 | 182 countries<br>(93%) | More external<br>investment enable<br>stronger immunisation<br>programmes<sup>3</sup> | GHED [27] |
| Health<br>financing –<br>private<br>expenditure | Private (e.g., Out<br>of Pocket<br>payments), PPP<br>per capita, mean<br>2015-2019 | 182 countries<br>(93%) | Increased private health<br>expenditure may<br>contribute to stronger<br>RI maintenance (though<br>a strong relationship is<br>less expected since RI<br>is often<br>government/donor<br>financed) | GHED [27] |
| Containment | First principal | 185 countries | Stringent containment | Oxford |
| policies | component from Principal Component Analysis (PCA) of eight containment indices mean annual score 2020-2022 | (95%) | policies, such as stay-at-home orders, may reduce perceived access to healthcare | COVID-19 Government Response Tracker (Ox-CGRT) [28] |
| Health policies (2 predictors) | First and second principal components from PCA of six health indices mean annual score 2020-2022 | 185 countries (95%) | Stronger containment policies to prevent and contain COVID-19 spread (e.g., mask mandates, testing protocols, vaccine eligibility) may reduce COVID-19 burden and thus health system overload | Ox-CGRT [28] |
| Economic policies | First principal component from Principal Component Analysis (PCA) of two economic indices mean annual score<br>2020-2022 | 185 countries (95%) | Economic support may facilitate adherence to stay-at-home measures and facilitate reduced COVID-19 caseload and health system burden | Ox-CGRT [28] |
| Mobility –<br>outside | First Principal<br>Component from<br>PCA of mean<br>annual change in<br>mobility compared<br>to 2019 baseline<br>for movement to<br>workplace,<br>grocery, retail,<br>transit, and parks<br>locations 2020-2022 | 127 countries<br>(65%) | Reductions in<br>movement outside may<br>indicate increased fear,<br>adherence to policies,<br>and be a proxy for<br>reduced healthcare<br>visits | Google [29] |
| Mobility –<br>residential | Percentage<br>change from 2019<br>baseline, mean<br>annual change for<br>2020-2022 | 127 countries<br>(65%) | Increased time at home<br>may indicate reduced<br>health seeking<br>behaviour | Google [29] |
| Country<br>wealth | Gross Domestic<br>Product (GDP)<br>PPP per capita,<br>mean 2015-2019 | 185 countries<br>(95%) | Richer countries able to<br>respond more<br>effectively to broad<br>pandemic challenges | World Bank<br>[30] |

### 2.2 Data

#### 2.2.1 Dependent variable

Our methodology for modelling RI coverage changes compared to previous temporal trends (in the absence of the pandemic) has been described previously [10]. In brief, we use Auto Regressive Integrated Moving Average forecasting to quantify the absolute difference, referred to as “coverage deltas”, between expected (ARIMA-modelled) and WUENIC-reported DTP3 coverage (based on latest data: July 2024 [11]).

#### 2.2.2 Independent variables

16 determinants were identified as relevant and available for inclusion. To enable cross-country comparison, indicators are quantified per capita (either pre-scaled, or by calculating based on United Nations World Population Prospects data [19]) and financial indicators (e.g., government health financing expenditure) are quantified in terms of Purchasing Power Parity (PPP^1^).

The final predictors are listed in **Table 1**, alongside hypotheses of potential impact. See Supplementary Materials section B for further details on data selection and cleaning.

After excluding incomplete data, the final dataset includes 119 countries and 356 country-year data points^4^.

#### 2.2.3 Principal Components Analysis (PCA)

To avoid over-fitting, where indicator sets have high internal correlation, we summarise by conducting centred and scaled PCA to reduce indicators to Principal Components (PCs) that capture > 50% of the variation. This applied to: eight containment indices (one PC capturing 64% variance), two economic policies (one PC covering 75% variance), six health policies (two PCs summarise 63% variance), and five outside mobility locations (one PC capturing 81% variance).

### 2.3 Univariate analyses

We explore the geographic variation and data available for each determinant by visualising on a global map. We explore pairwise relationships between each determinant and coverage deltas by plotting scatterplots with Loess regression [31]. See Supplementary Materials section B and C for these analyses.

### 2.4 Multivariate analyses

We employ a mixed-methods approach to identify the factors associated with changes in RI during the pandemic. First, we conduct stepwise linear regression, allowing for straightforward interpretability of the coefficients. Secondly, Random Forest regression to identify potential nonlinear interactions, complex patterns, or collinear effects that linear regression might not detect.

#### 2.4.1 Linear regression

Linear regression models were built incrementally, adding explanatory variables according to the causal inference framework (**Figure 1**). By selecting the stepwise order, we were able to factor in the conceptual framework and pre-described relationship between variables and reduce the risk of overfitting which may occur using automated methods.

Independent variables were iteratively added and the relationship between the current best fitting model residuals and the newly added variable was reviewed visually to check the existence of a non-linear relationship. Where non-linear we create a synthetic “threshold” binary indicator to flag the turning point (coded 0 for below the threshold and 1 after) and then construct a new model including the interaction term. Predictors were selected for inclusion by evaluating (i) the *p*-value associated with each variable’s *t*-statistic, and (ii) the Bayesian Information Criterion (BIC) of consecutive models with the objective of only including variables with strong association and explanatory power. Correspondingly, we retain variables where (i) *p* < 0.001 to minimise the probability of including independent variables in the final model by chance, and (ii) the BIC decreases with variable addition, to factor-in the trade-off between explanatory power and parsimony.

#### 2.4.2 Random Forest

We randomly split the dataset into training (70%) and testing (30%) subsets. We build a Random Forest with 500 trees, each tree considering 5 variables at each split to introduce diversity and prevent overfitting, using the ‘*caret*’ package in R [32]. Variable importance is assessed by (i) the Mean Decrease in Accuracy (MDA) when the predictor is excluded, and (ii) the Mean Decrease in Impurity (MDI) which represents feature importance across all trees. Importance score ranking is compared to linear regression results. We evaluate the overall explanatory power of the Random Forest by calculating the R-squared of the resulting Random Forest model using the predictor weightings applied to the test dataset.

## 3. Results

### 3.1 Linear regression

The best fitting model describes 20% (adjusted *R*^2^ = 0.196, F-statistic: 22.57, and *df* = 351) of the variation in observed pandemic coverage deltas. This model includes two explanatory variables: pre-pandemic immunisation programme strength and health workforce capacity. Both predictors exhibit non-linear relationships, thus include synthetic binary variables to describe the respective turning points. The final linear model results are summarised in **Table 2**. After accounting for pre-pandemic immunisation performance and health workforce capacity, none of the remaining predictors had sufficiently strong evidence for inclusion – see Supplementary Materials section C for full stepwise linear modelling results.

**Table 2.**
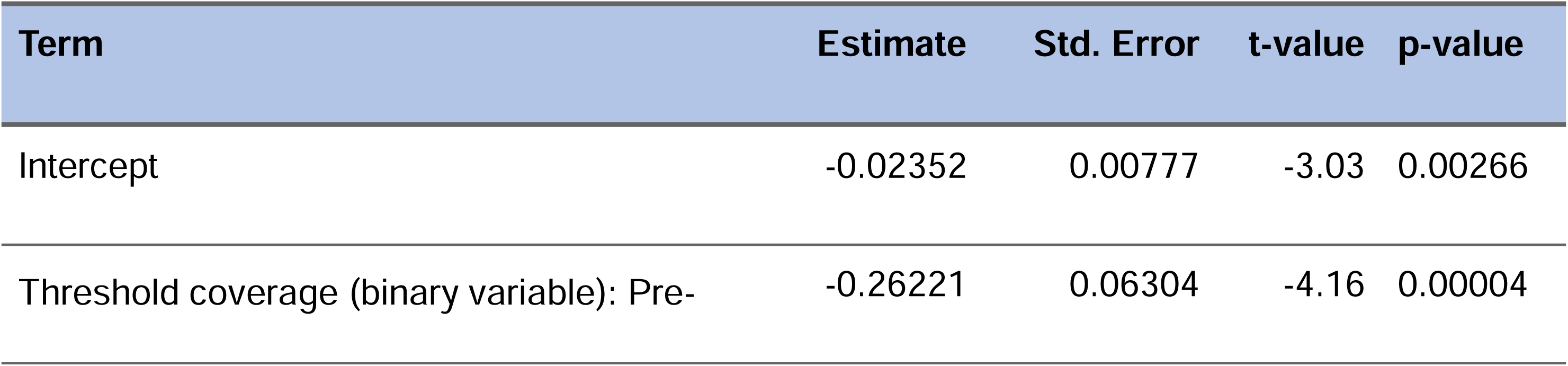
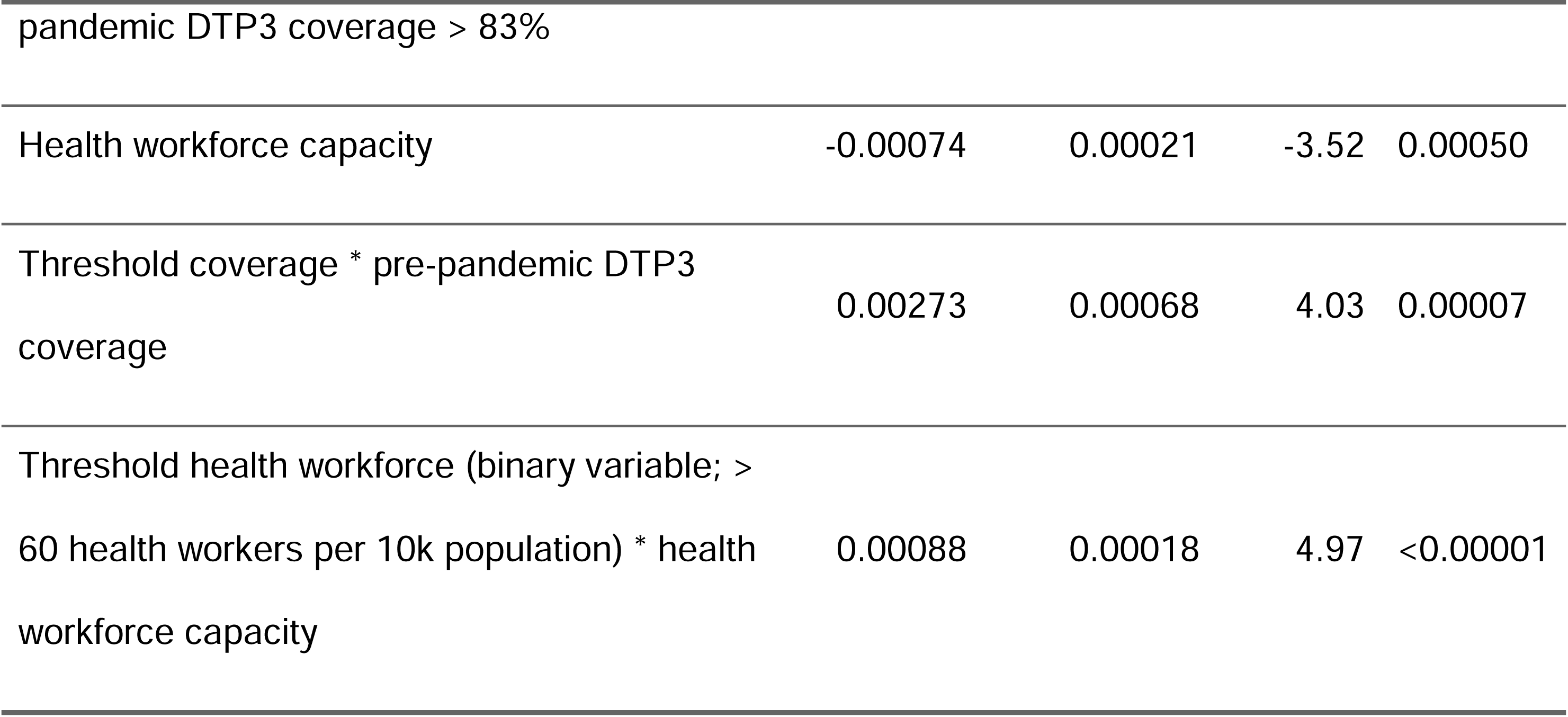
Summary results from the best fitting model identified through stepwise regression.

Pre-pandemic immunisation programme strength provides limited explanatory power below the threshold of ∼83% pre-pandemic DTP3 coverage (only the interaction terms are included in the final model). Beyond this threshold, higher pre-pandemic DTP3 coverage is associated with better performance during the pandemic. After adjusting for immunisation programme strength, up until ∼60 health workers (doctors and nurses) per 10,000 population, coverage deltas worsen with increasing number of health workers^5^. After this, access to more health workers is associated with better maintenance of RI performance during the pandemic.

### 3.2 Random Forest

The *R*^2^ of the Random Forest on the training dataset is 38.1% and 10.0% on the test dataset. Much of the variation in coverage deltas is not well-described by the Random Forest: *R*^2^ values are low, and the model appears to be overfitting since the training data details do not generalise to the test data.

Random Forest importance scores support the linear regression model findings – identifying pre-pandemic immunisation programme strength (MDA: 15.88%; MDI: 0.09%) and health workforce capacity (MDA: 15.64%; MDI: 0.08%) as the two most important predictors. Six additional variables have comparatively high importance values (over 10%) indicating additional predictors, potential correlation, or more complex interactions. These additional variables are broader health systems strength (MDA: 12.25%, MDI: 0.04%), and financing indicators: government health financing expenditure (MDA: 10.84%, MDI = 0.04%), GDP (MDA: 10.61%, MDI: 0.05%), and private health financing expenditure (MDA: 10.43%, MDI: 0.08%). See **Figure 2** for a full summary of importance scores per predictor.

**Figure 2.**
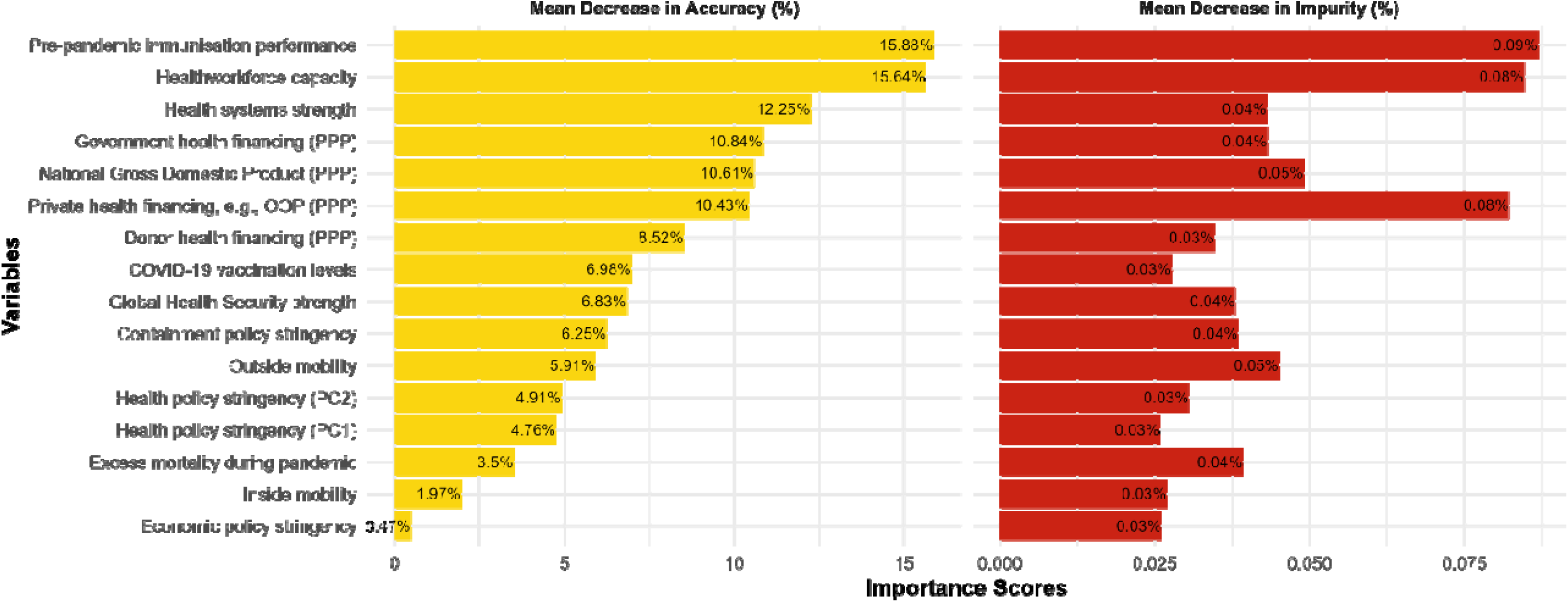
Variable importance scores from the Random Forest regression. The Mean Decrease in Accuracy (MDA) is calculated by permuting the values of each variable and measuring the resulting decrease in the model’s accuracy. Mean Decrease in Impurity (MDI) measures the total reduction in impurity (e.g., Gini impurity) brought by a variable across all trees in the forest. It reflects the variable’s ability to split the data into more homogeneous nodes. Both MDA and MDI scores are relative to a given model, and higher values indicate that a variable is more important (MDA) and is better splitting the data effectively (MDI).

## 4. Discussion

We provide evidence (*p* < 0.001) of parabolic relationships between both pre-pandemic immunisation programme strength and health workforce capacity and the outcome of pandemic RI coverage resilience. The turning points are about 83% coverage and approximately 60 health workers per 10,000 population respectively. Health systems strength and multiple financial indicators (GDP, private and government health financing) are also important predictors of pandemic RI performance, based on the Random Forest. 80% of the variation in pandemic RI coverage performance remains unexplained after exploring the contributions of 16 variables covering health systems, pandemic policy, mobility, and health indicators.

### 4.1 Interpretation of models

It is encouraging to quantitatively demonstrate resilience associated with robust immunisation infrastructures. Below ∼83% coverage – slightly below the global average DTP3 coverage of ∼87% – pre-pandemic immunisation provides limited explanatory power. This may be driven by poorer data quality in countries with weaker immunisation systems obscuring potential relationships or confounding from other factors. Alternatively, a minimum threshold in programme performance may be essential to safeguard against disruptions – with this coverage indicating a sufficient programme expertise and management foundation.

It is equally reassuring to identify a positive relationship between health workforce capacity and pandemic RI performance above the threshold. This is noteworthy as the relationship between absolute immunisation coverage and health workforce density (measured by nurses, midwives and community health workers, CHWs) has previously been found to be weak [12]. Below the threshold, the negative relationship requires careful interpretation. Our reliance on only doctor and nurse data, due to data quality and availability, may obscure identification of trends, since immunisation systems are often staffed by other crucial health worker cadres including CHWs and volunteer health officers [33]. Use of national averages may hide important density distribution insights with greater needs in fragile areas or with dispersed populations.

Additional predictors identified via the Random Forest regression – health systems strength and financial indicators – likely reflect the underlying contribution of financial investment in broader health systems to achieving immunisation-specific outcomes (and corresponding high internal correlation of these indicators with other included variables). This supports previous research – economic characteristics such as GDP were found to be important determinants of immunisation coverage over the 41 years pre-pandemic [13].

### 4.2 Other predictors

We found limited or no evidence of association between other predictors and coverage deltas.

This included COVID-19 vaccination coverage, suggesting that COVID-19 vaccine campaigns did not detract from RI provision, as previously reported [3] – suggesting catch-up may have occurred after temporary disruption. There was however no evidence of synergies such that COVID-19 rollout strengthened RI, as hoped through campaign synergies.[3]

Fear of contracting COVID-19 was often cited by parents as reasons for delaying or not seeking RI for their children [17]. We found some hints of a trend that excess mortality (as a proxy for COVID-19 direct impact) may be associated with decreases in RI coverage. Risk perception during COVID-19 varied across countries, correlated with experiences of the virus and socio-cultural factors [34] – thus the insufficient evidence to include this variable in the final model does not necessarily contradict qualitative reports.

There was also a potential trend such that stronger reductions in outdoors mobility related to reduced pandemic RI performance. We lack specific data on movement to health facilities, and mobility outdoors may poorly represent health seeking behaviours. This data unrepresentativeness for our purposes seems probable given lockdowns, lack of transport, and health facility closures are all key drivers of reported reduced RI through parental and health worker surveys [17].

The absence of predictive power from pandemic policies suggests, positively, these restrictions did not interrupt routine service provision. It may also reflect dependence on policy adherence and effectiveness – with populations following restrictions to different extents over time [35], and with sub-national variation.

### 4.3 Strengths

Our methodology is transparent and reproducible, leveraging publicly available datasets and shared R scripts [36], and could be easily adapted to other contexts or health areas.

Employing a mixed method of linear modelling and Random Forest regression is valuable because it leverages the strengths of both traditional statistical and modern machine learning approaches, offering a comprehensive analysis.

### 4.4 Limitations

Country-level research using national averages may hide important inequalities and inequities in RI disruption and impact. We welcome sub-national analyses, to uncover geographic disparities that may inform micro-planning exercises, and help guide effective, tailored, and resource-efficient strategies to catch-up missed children.

Our analyses were limited to available, nationally-summarised data, and exclude additional indicators that may help explain RI performance – particularly vaccine hesitancy, given anti-vaccine sentiment during the pandemic. We explored publicly available vaccine hesitancy datasets [37,38], but determined insufficient completeness (covering < 80, primarily high-income countries) and challenges generating national representative, comparable values over different survey years and sub-populations.

Due to data incompleteness, approximately 75 countries – mainly in Africa and Asia – were excluded. Our results may therefore not well represent low- and middle-income countries.

### 4.5 Areas for future research

Further research could explore which immunisation service provision components are key for buffering disruptions, e.g., outreach services or health communication. Identification of health worker cadres most crucial to RI could help highlight gaps in hiring, and flag priorities for retention and training. Exploration of the role of additional predictors is welcomed.

## 5. Conclusion

Our findings underscore the role of robust immunisation programmes and sufficiently sized health workforces in mitigating RI disruption during global health crises once above minimum thresholds (about 83% DTP3 coverage and 60 health workers per 10,000 population). Financial resources (particularly government and private health expenditure and GDP) and broader health systems strength also impact RI disruption. However, most of the variation in DTP3 coverage deltas is not explained by our predictors: 20% was explained through linear regression and 10-38% through Random Forest regression.

We did not find evidence that pandemic policies, changes in mobility, COVID-19 vaccination, or COVID-19 burden had important effects on pandemic RI coverage. This was unexpected given the extensive global focus on many of these areas during the pandemic. This may reflect the bi-directional and multi-faceted effects of these interventions, or data challenges.

We encourage continued efforts to identify RI disruption determinants. We hope for increased dataset availability that would enable more granular analyses and facilitate more specific programmatic recommendations to build resilient immunisation programmes.

## Supporting information

Guideline checklist

## Data Availability

All datasets are publicly available in our Git Hub repository alongside the scripts for all analyses, which were conducted in R.

https://github.com/bevans249/pandemic_RI_coverage_determinants

## Acknowledgements

BE is a PhD student at the Institute of Global Health, Faculty of Medicine, University of Geneva, under supervision from LK, and received no funding. OK was supported by the Swiss National Science Foundation Grant [grant number PP00P3_202660]. TJ was supported by the MRC Centre for Global Infectious Disease Analysis [grant number MR/R015600/1], jointly funded by the UK Medical Research Council (MRC) and the UK Foreign, Commonwealth & Development Office (FCDO), under the MRC/FCDO Concordat agreement and is also part of the EDCTP2 programme supported by the European Union. These funders had no role in the design and conduct of this study; collection, management, analysis, and interpretation of the data; preparation, review, or approval of the manuscript; and decision to submit the manuscript for publication.

## Footnotes

1 which converts different currencies and weights for costs of goods in a country to enable comparisons across countries

2 Percentages are report in relation to the 195 countries included in WUENIC data

3 Noting that external health expenditure likely substitutes for government financing in low- and middle-income contexts

4 Vietnam is only included for 2020 and 2021, due to missing data in 2022

5 This threshold is higher than the univariate analysis turning point, and is based on visual assessment of the residuals

