## Supplementary Materials for "Pre-pandemic national immunisation programme strength and health workforce capacity improved routine immunisation resilience during the COVID-19 pandemic"

Contents

A. Conceptual framework
B. Data sources: selection and cleaning
C. Univariate analyses
D. Stepwise linear regression results
E. Reproducibility
A. **Conceptual framework**

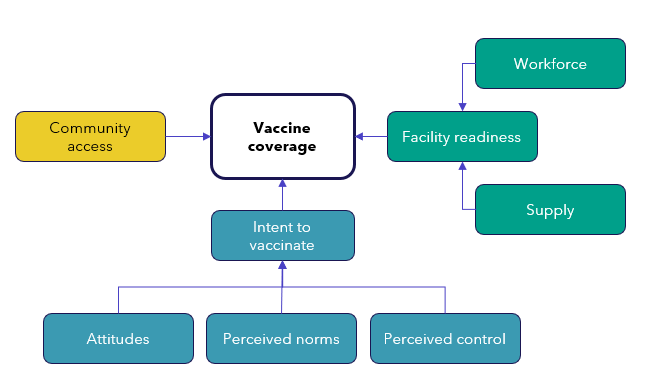
The original conceptual high-level conceptual framework, developed by Philips et al., [1] and adapted for this research is depicted in **Figure 1**.

**Figure 1** the Conceptual framework for determinants of vaccine coverage in low- and middle-income countries, developed by and reproduced from Phillips et al., 2017. Three dimensions of determinants are identified: access to services (yellow), facility readiness in terms of supply and workforce availability and quality (green), and user intention to vaccinate (blue).

B. **Data sources and selection**

Pre-pandemic immunisation programme performance

It is hypothesised that countries with well-established robust immunisation systems that routinely achieve high coverage will have been able to maintain or recover immunisation performance during the pandemic, as a result of a pre-existing strong Extended Programme on Immunisation (EPI) with good programme management and previous experiencing buffering adverse events. We use DTP3 as a proxy indicator of performance across countries, as it is used as a key tracer for immunisation strength across countries and time [2].

We calculate mean DTP3 coverage for 2015-2019 (excluding missed values) which reports vaccination coverage for children aged under one-year-old. This indicator is sourced from WHO and UNICEF Estimated of Immunisation Coverage (WUENIC) based on the latest available dataset from July 2024 [3], which includes retrospective updates to previous estimates e.g., due to later validation of new surveys. WUENIC data is published with a transparent methodology that factors in administration data and population-level representative surveys [4,5].

We visualise the underlying variation in country performance from this dataset in **Figure 2**.

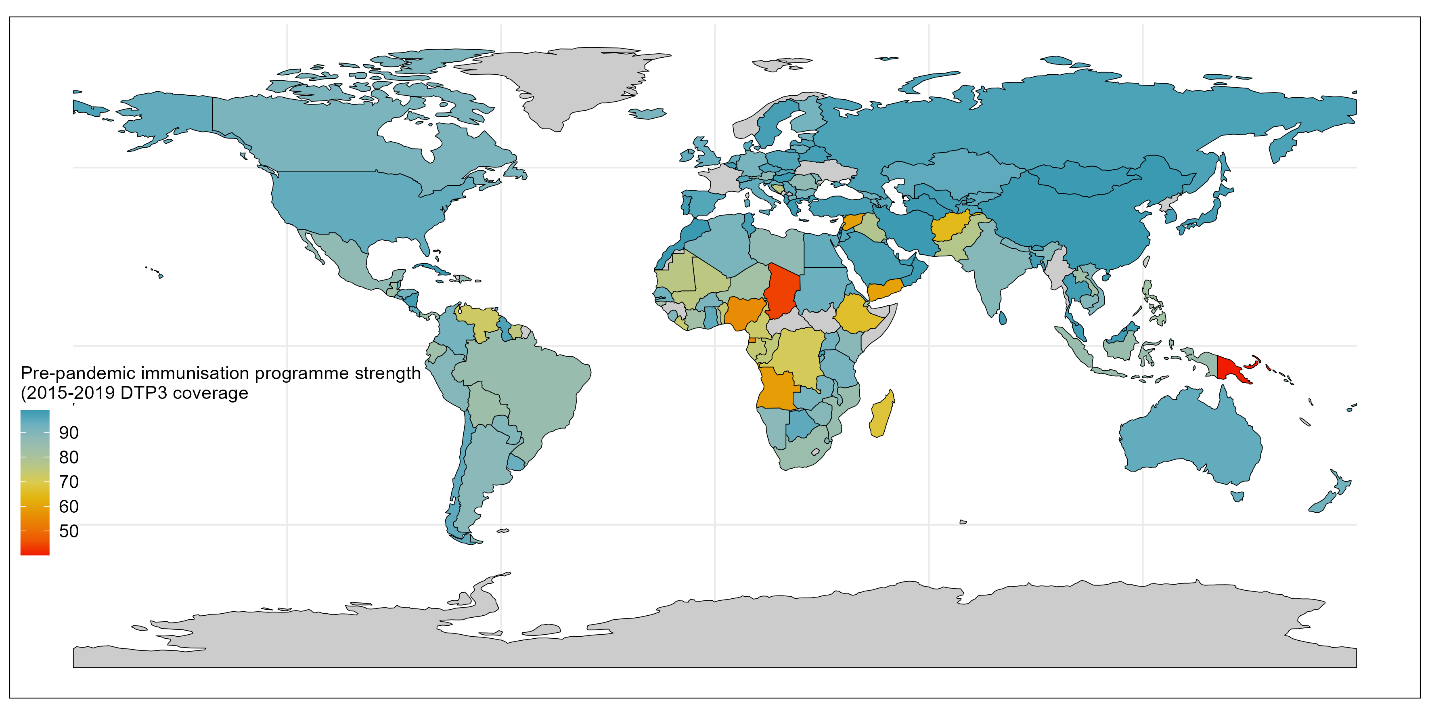

**Figure 2** Pre-pandemic immunisation programme strength variation globally, as measured by 2015-2019 mean DTP3 coverage. Colour coding indicates the programmes strength, with red signalling poor programmes and bluer signalling strong programmes. Grey areas indicate states without data for this indicator.

Health workforce capacity

Health workers – quantity thereof, and capacity – are essential to the provision of health services. The term ‘health workforce’ is expansive and considers "all people engaged in actions whose primary intent is to enhance positive health outcomes" [6]. Unfortunately, datasets on key cadres of health workers such as volunteer health workers or community health officers, are non-existent or poor. As a result, for this analysis we use data on doctors and nurses as a proxy for broader health workforce.

We use WHO datasets for the number of doctors and nurses per 10,000 people per country, calculating the mean of the reporting cohorts for 2015-2019 (excluding missing values) [7]. After investigating the relationship between each of doctors and nurses and pandemic RI coverage we summed the number of doctors and nurses for model fitting, since both relationships exhibited the same trends.

We visualise the underlying variation in country health workforce capacity from this dataset in **Figure 3**.

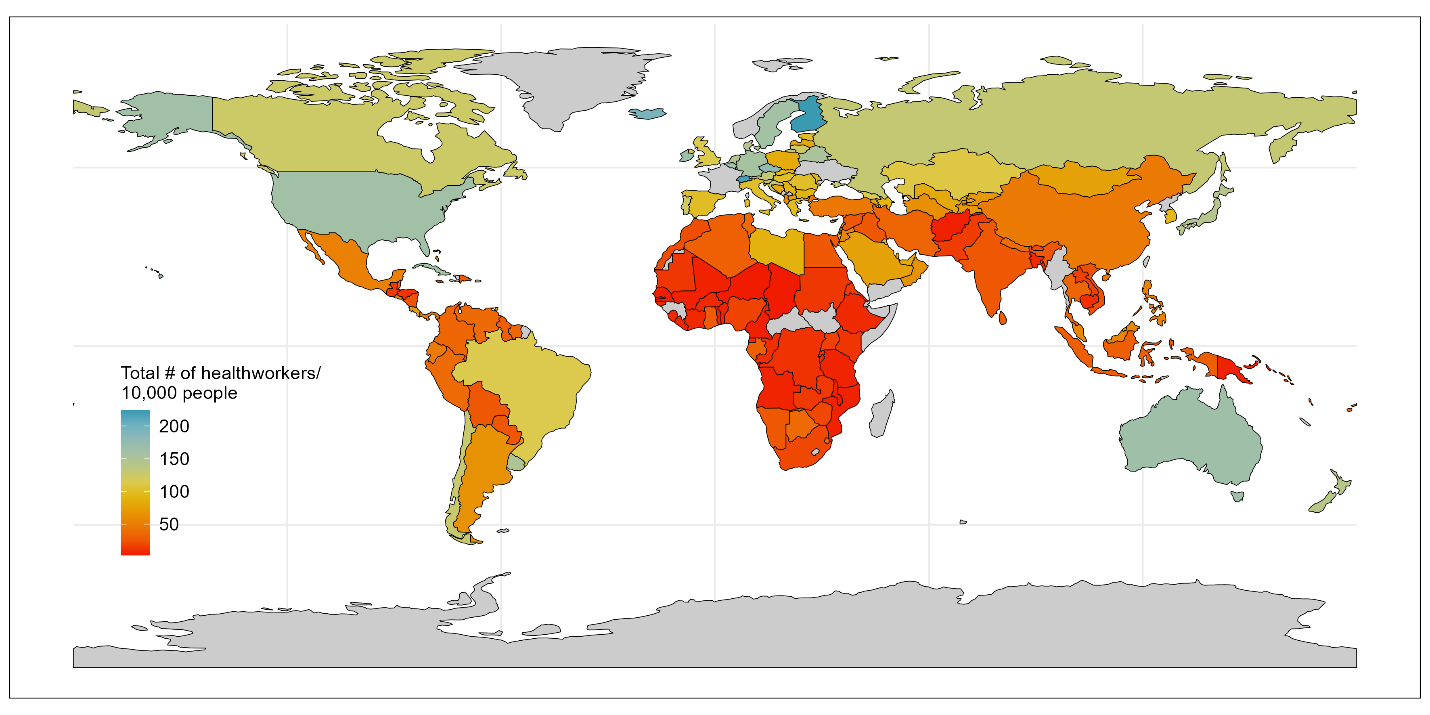

**Figure 3** Health workforce capacity variation globally, as quantified by mean number of doctors and nurses per 10,000 population from 2015-2019. Colour coding indicates the relative number of health workers per capita, with red signalling less workers than blue. Grey areas indicate states without data for this indicator.

Health system strength

Health systems strengthening is defined by the WHO as “the process of identifying and implementing the changes in policy and practice in a country's health system, so that the country can respond better to its health and health system challenges”, thus by definition, a stronger health system would be expected to be able to respond better to a pandemic and maintain core health services.

Quantification and comparison of health system strength across countries requires a clear understanding of the goals and functions of a health system and a framework with quantifiable underlying metrics. The WHO’s health system “building blocks” [8] are often used for this purpose, since they offer a relatively simple definition of a health system with six pillars – (i) financing, (ii) health workforce, (iii) information systems, (iv) medical products and technologies, (v) leadership/governance, and (vi) service delivery – and 17 recommended core indicators for their quantification. A limitation in direct use of the WHO framework lies in the lack of available (and publicly accessible) datasets to quantify the 17 core indicators – or even a proxy indicator per building block.

As a result, we sought to identify an alternative dataset that would describe the *outputs* of health system strength, rather than the *inputs*. The Universal Health Coverage Index (UHCI, [9]) aims to provide a summary measure of health service coverage based on 14 tracer indicators across four areas – (i) reproductive, maternal, newborn and child health, (ii) infectious diseases, (iii) non-communicable diseases, and (4) service capacity and access. Whilst it is noted by the WHO that there are some gaps in data availability, this indicator appeared to be the best proxy for what the WHO health system building blocks define as the aims of “access” and “coverage”. The use of this indicator also aligns with other global analyses, since the UHCI is used for monitoring progress against the Sustainable Development Goal (3.8.1). This indicator is reported as a index, reflecting breadth of UHCI coverage across the total country population.

Quantification-wise, the index scores country from 0-100 and is available for odd numbered years. We calculate the mean UHCI from 2015-2019 from the three available years in that time period (excluding missing values).

We visualise the underlying variation in country performance from this dataset in **Figure 4**.

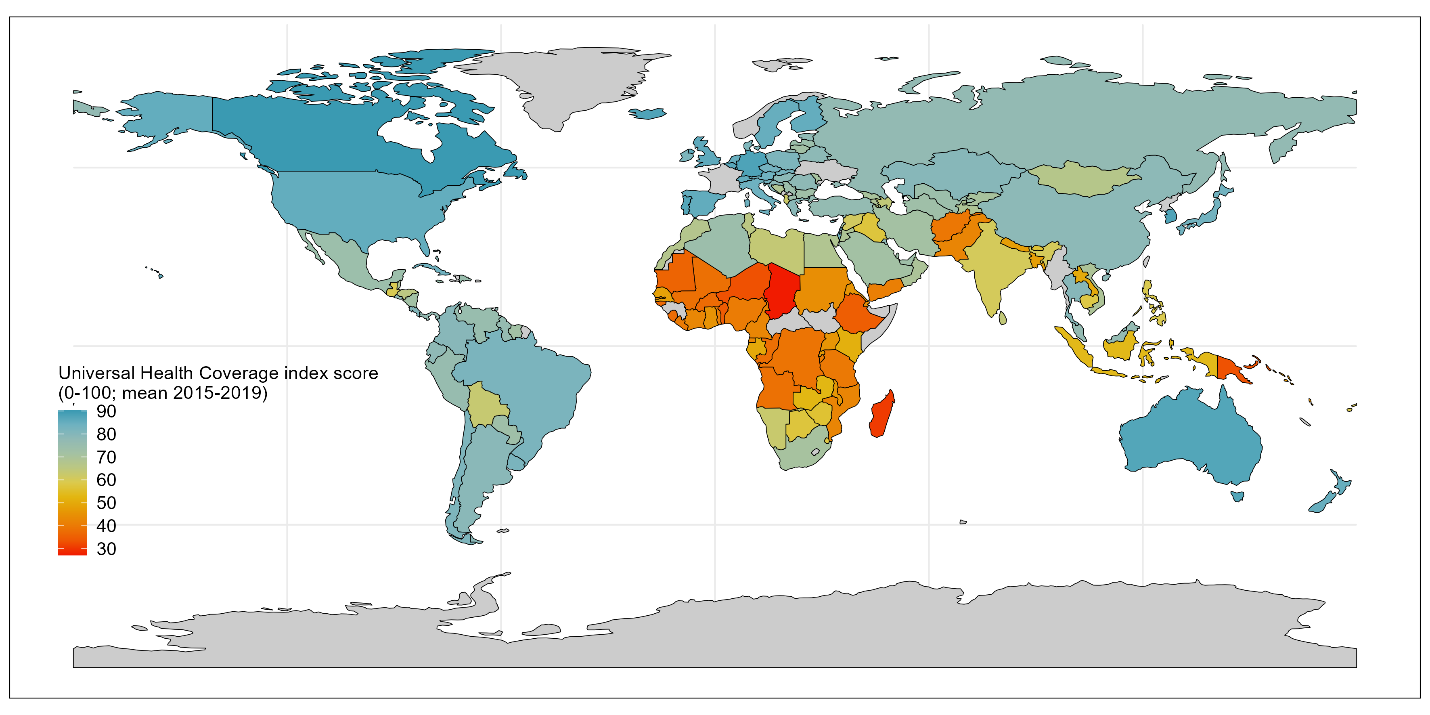

**Figure 4** Health system strength variation globally, as quantified by Universal Health Coverage Index (UHCI) mean score (2015-2019). Colour coding indicates the relative health system strength across countries, with red signalling low UHCI scores and blue signalling high scores and strong health systems. Grey areas indicate states without data for this indicator.

Global health security

Global health security (GHS) is defined by the World Health Organisation as “the activities required, both proactive and reactive, to minimize the danger and impact of acute public health events that endanger people’s health across geographical regions and international boundaries” [10]. It thus follows that countries with better GHS are intended to be able to prevent and respond to threats including pandemics. We include GHS index quantified pre-pandemic, in 2019, to investigate whether the hypothesised association holds that countries with higher GHS are able to better maintain routine immunisation services during the pandemic and suffer less reductions in coverage.

The Global Health Security Index (GHSI, [11]) is the first and only publicly available dataset that aims to quantify the relative readiness of countries to respond to pandemics and other global emergencies. It is comprised of 171 questions grouped into 37 indicators across six categories, all compiled from publicly available information, with indicators selected based on “literature review, and standard accepted measurements for global health security as captured in the International Health Regulations Joint External Evaluation tool and elsewhere” [12]. The GHSI is available for 2019 and 2021. For consistency with other indicators, we use the pre-pandemic (2019) values), however we use the version of these calculated by the methodology updated in 2021 which retrospectively re-scored the GHSI for 2019 based on inclusion of additional indications and changes to weighting of indicators.

In addition, it is noted that some of the indicators within the GHSI draw on sub-indicators that are included or closely related to other variables quantified separately in this analysis, e.g., pre-pandemic immunisation coverage and number of doctors and nurses. Since these overlaps represent only a handful of the 171 input datapoints to the GHSI, the GHSI was not recalculated to exclude these variables as the impact of duplication is foreseen as minor and comparable across all countries.

We visualise the underlying variation in country performance from this dataset in **Figure 5**.

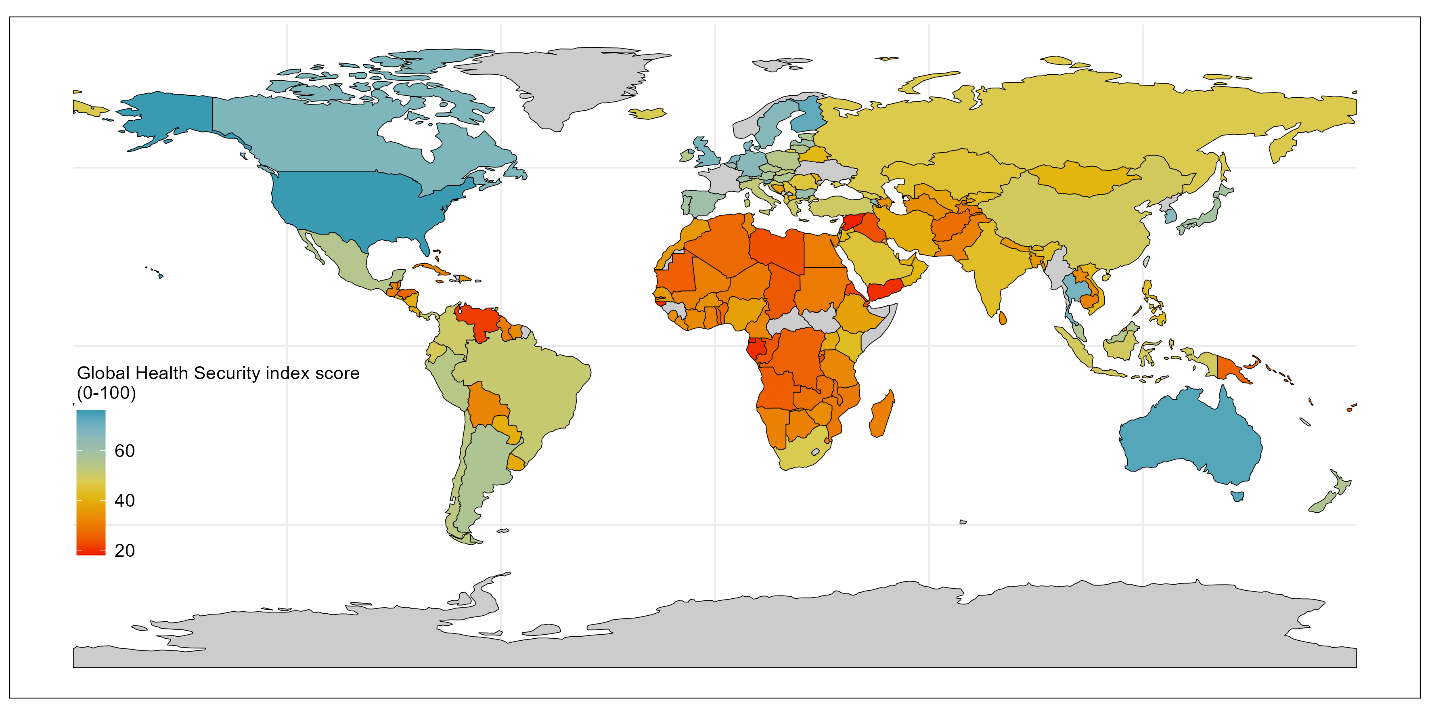

**Figure 5** Global health security globally, as quantified by the Global Health Security index (GHSI; from 2019). Colour coding indicates the relative strength of pandemic preparedness across countries, with red signalling low GHSI scores and blue signalling high scores and better preparedness. Grey areas indicate states without data for this indicator.

COVID-19 vaccination

COVID-19 vaccines were developed, tested, and approved for emergency use within 11-months of the declaration of the pandemic. Despite calls for global equitable access to COVID-19 vaccines during the acute phases of the pandemic, access to vaccines varied considerably – with high-income countries securing the majority of vaccines, and the COVAX mechanism supporting access to lower-income countries. When vaccines were available, COVID-19 vaccination campaigns required significant resources due to their scale. We wished to explore whether focus on COVID-19 vaccination may have come at the expense of focused efforts on infant immunisation – as reported in WHO Pulse Surveys [13], or whether COVID-19 vaccination offered opportunities for RI integration and catch-up.

To explore this potential association, we calculated the number of COVID-19 vaccinations administered per 100,000 (total) population per year, by scaling absolute vaccination administration data from Our World In Data using the UN WPP total population estimates per year from 2020-2022 [14]. Where no data is reported we assume this reflects zero vaccines administered and relates primarily to some countries in 2020 when vaccines were initially rolling out globally.

We visualise the underlying variation in country COVID-19 vaccination levels from this dataset in **Figure 6**.

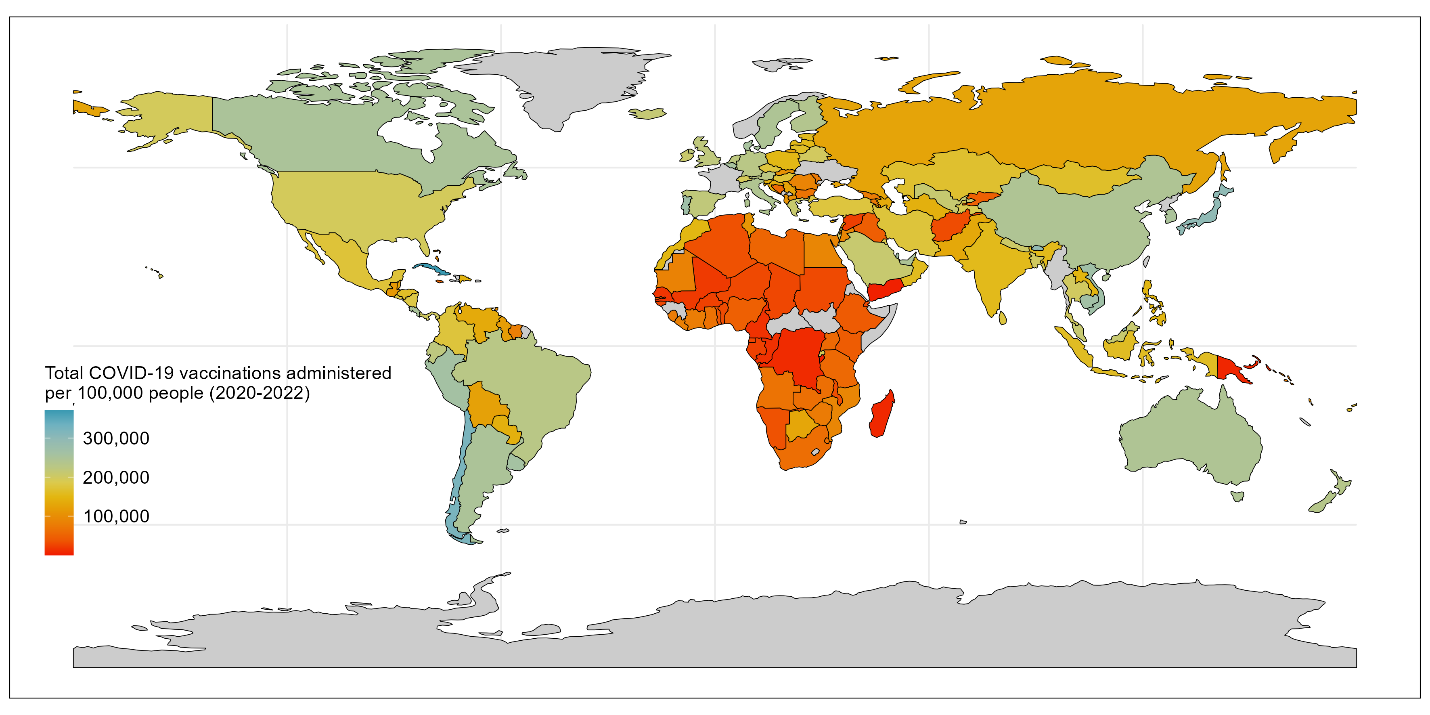

**Figure 6** Number of COVID-19 vaccines administered from 2020-2022 per 100,000 population. Colour coding indicates the extent of COVID-19 vaccination across countries, with red signalling low reach and blue signalling high coverage. Grey areas indicate states without data for this indicator.

COVID-19 burden

During the peak period of the pandemic, countries with high burdens of COVID-19 cases reported that their health systems were overwhelmed [13,15,16] – with acute care hospitalised beds fully utilised and large numbers of patients requiring intensive care and oxygen supplies. Some countries cancelled routine surgery in order to offer increased hospital capacity to pandemic response. Some countries constructed emergency hospitals at record speed to increase bed capacity further. These efforts diverted human and physical resources from other routine health service provision to COVID-19, and it therefore hypothesised that countries with a higher severe case burden would need to divert more resources to COVID-19 and that routine immunisation may have suffered more in these countries. Increase COVID-19 burden may also have led to increased fear of contracting the disease in the population and contributed to changed behaviours and less health seeking behaviour (as reported by parents across many countries [17]).

Comparing country-level COVID-19 case and death data is challenging due to varied access to COVID-19 diagnostics, differing testing policies, and variations in death reporting guidelines (i.e., what counts as a COVID-19 death), as well as poor data availability for both morbidity and mortality data in many countries. As a result, rather than include poor data with large gaps, we sought to include a proxy indicator for COVID-19 health impact in the form of modelled excess mortality estimates. To our knowledge, four sources have compiled estimates of excess mortality for all countries for at least 2020 and 2021 – the World Mortality Dataset [18], the WHO [19], the Economist [20], and the COVID-19 Excess mortality Collaborators [21]. The World Mortality Dataset currently provides data for 127 countries and territories, and therefore we preferred alternative, more complete, datasets. Whilst the WHO provides estimates for 194 countries, and the World Mortality Dataset covers 191 countries and territories, however both only covers the period from 2020-2021 and since we intend to conduct analyses over a three-year period (2020-2022) we opted to use the Economist data which covers 223 countries and regions and covers 2020-2023.

Economist excess mortality data includes a weekly updated time series, including an indicator for ‘cumulative excess mortality per 100,000 population’ [22]. We extract the end-of-year values for this indicator for 2020, 2021, and 2022 and then back calculate annual excess mortality for 2021 and 2022 as the difference between the end year and previous end year values.

We visualise the underlying variation in country excess mortality from this dataset in **Figure 7**.

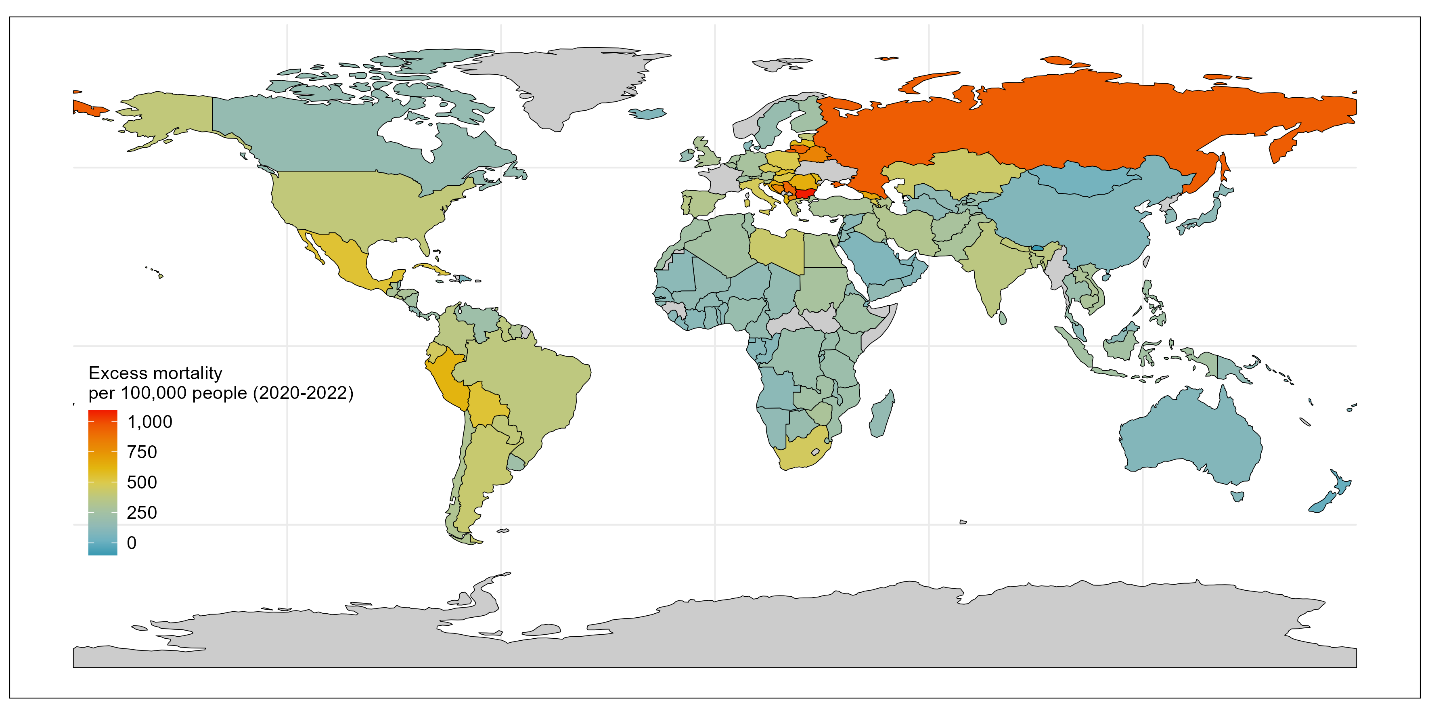

**Figure 7** Excess mortality per 100,000 population (2020-2022), as a proxy for COVID-19 burden. Colour coding indicates the extent of COVID-19 burden, with red signalling high excess mortality and blue signalling minimal additional deaths than expected during the pandemic. Grey areas indicate states without data for this indicator.

Pandemic policies: containment, economic, and health

During the COVID-19 pandemic, unprecedented policies were introduced to help prevent cases, identify outbreaks, and protect vulnerable populations (amongst other goals) [23]. We hypothesised there may not be as strong a relationship between pandemic policies and DTP3 coverage deltas since pandemic policy stringency is moderated by adherence to polices, and outcome measures – such as COVID-19 burden, COVID-19 vaccination level, and changes in movement and behaviour – may capture variation partly stemming from policies. Nevertheless, there could be residual variation driven by variation in pandemic policy stringency, with more stringent containment policies reducing mobility and health seeking behaviour; more stringent economic policies enabling increased adherence to stay-at-home orders and thus reducing COVID-19 transmission and enabling more “business as usual” including routine health services; and stronger health policies also reducing COVID-19 burden and thus facilitating continuation of routine services.

Country national, regional, and local, policy responses during the pandemic were collated and codified in the Oxford Covid-19 Government Response Tracker (OxCGRT) on a daily basis from 2020-2022 [24]. Eight containment policy categories (covering restrictions on movement and gatherings), four economic policy categories (covering financial support to reduce financial burden), and eight health policy categories (covering diagnostics, vaccination, and mask wearing amongst other policies) were recorded and codified from 0 to up-to 4, with higher numbers indicating greater stringency of policies. In some cases, multiple versions of each indicator exist with variations signalling whether the policy applied to all populations, the majority of the population, vaccinated people only, or non-vaccinated individuals at a national or sub-national level. These individual variables were also published summarised into five indices (one depreciated) – with overlapping inclusion of indicators within each index.

Our focus is on national-level broad trends; therefore, we focus on national-level indicators applied to most of the population. First, we removed four indicators that were flagged by the authors as incomplete and not updated (two economic and two health indicators: E3, E4, H4, and H5) [25]. We then sub-selected the version of all remaining indicators (eight containment, six health, and two economic) that provides a stringency for policies as applied to most of the population at a national level for each year from 2020 to 2022 inclusive. We calculated an annual mean stringency score per year for each of these 16 indicators.

To reach a manageable and meaningful summary set of policy variables, we then used Principal Components Analysis (PCA) to reduce each category of data into Principal Components that described at least 50% of the underlying variation per sub-category. For the containment policies the Principal Component described 64% of the variation in the data and is composed of a relatively similar proportion of information across each of the underlying eight variables (8-15% of each). We thus kept a single Principal Component. For the two economic indices the Principal Component described 75% of the variation in the data and is composed of equal amounts of each variable, and thus again we kept a single Principal Component. For the health indicators the diagnostic testing policy and public information campaigns contributed the most (together explaining ~50% of the variation) followed by vaccination policy and mask wearing (explaining an additional ~40%), with minimal contribution from contract tracing or elderly protection. We maintained two health policy variables for fitting in the model, so as to capture > 50% of the variance from the underlying policies.

After conducting PCA, pandemic policy Principal Components are all scored such that higher scores represent more stringent policies, e.g., stronger containment policies (such as more frequent or stringent school or workplace closures), more health policies (such as requirements to wear masks more often), and more economic support (e.g., broader economic relief during the pandemic).

We visualise the underlying variation in country policy stringency from this dataset in **Figure 8** (containment policy), **Figure 9** (economic policy) and **Figure 10** (health policy).

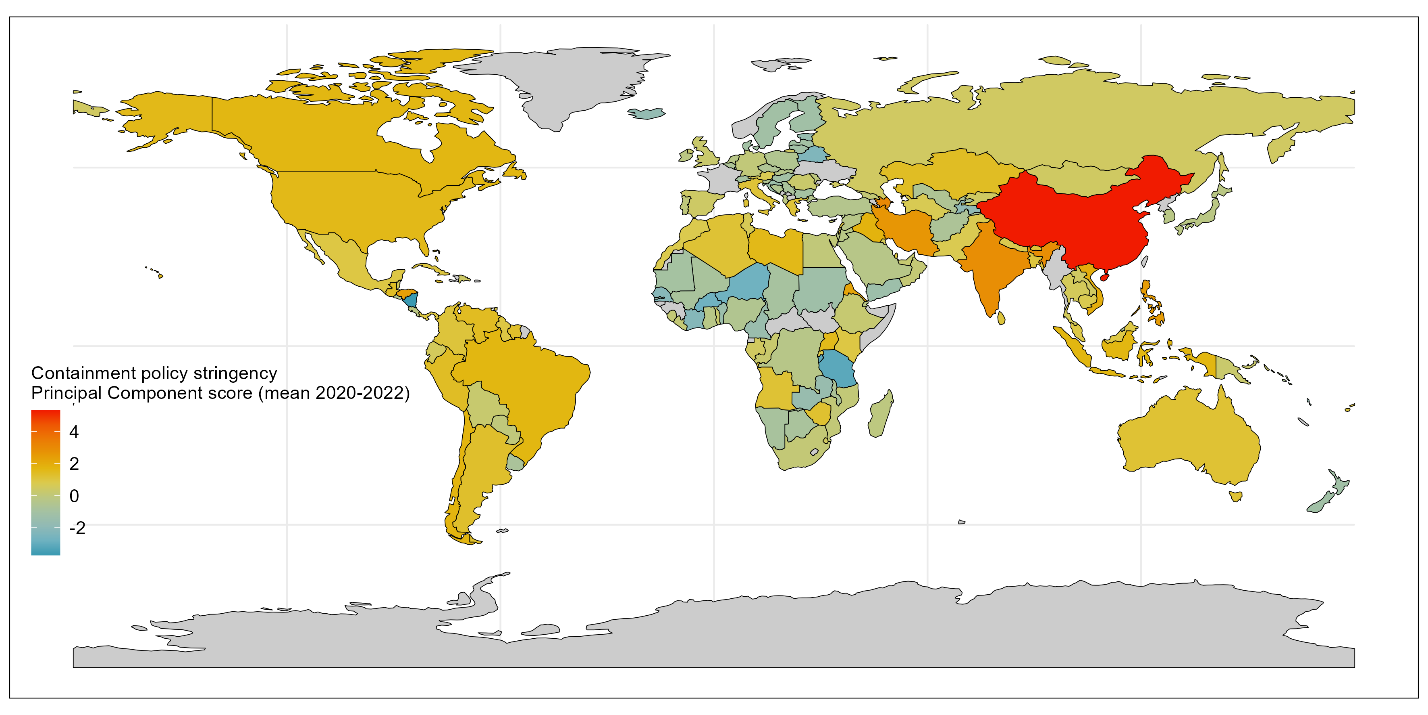

**Figure 8** Containment policy stringency variation across countries, as quantified by a single Principal Component summarizing 6 containment policies. Higher stringency values indicate more stringent containment policies such as more frequent or stringent school or workplace closures. Colour coding indicates the policy stringency, with red signalling more restrictions and blue signalling fewer restrictions on average during the pandemic. Grey areas indicate states without data for this indicator.

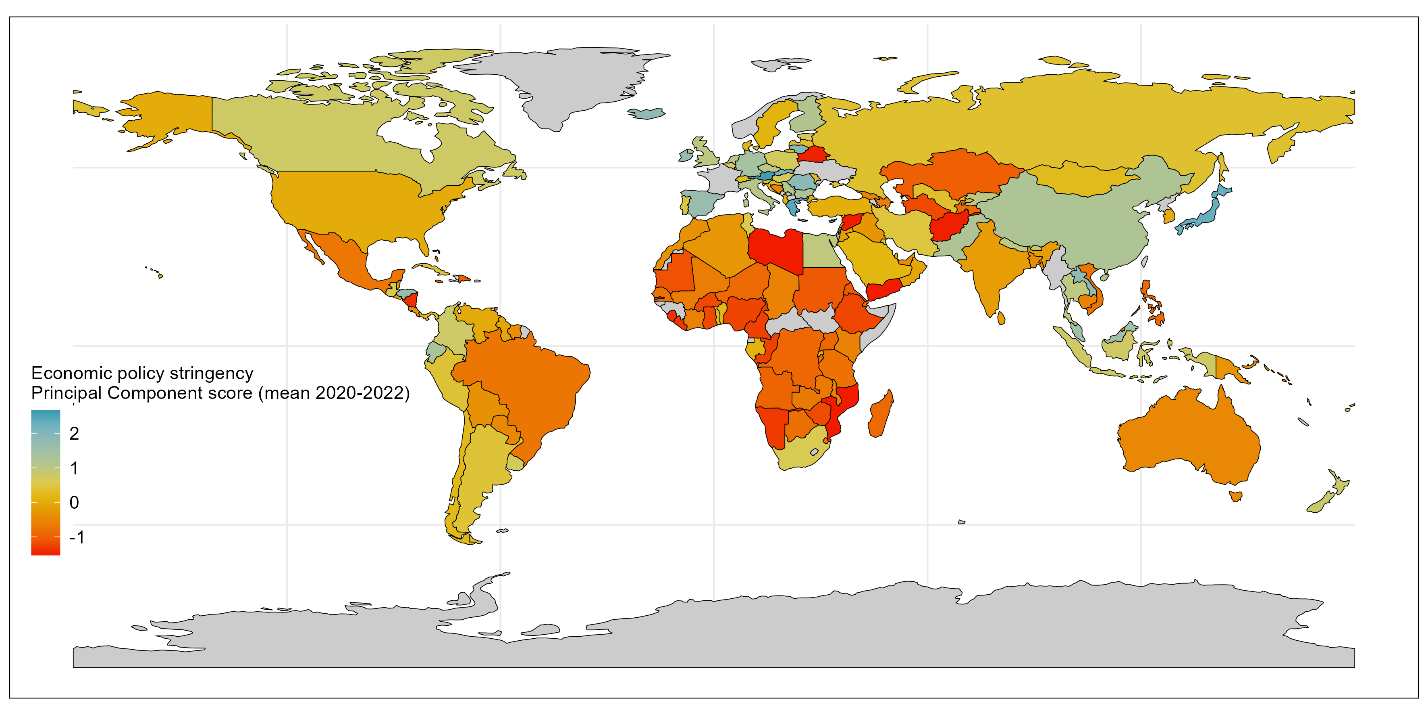

**Figure 9** Economic policy stringency variation across countries, as quantified by a single Principal Component summarizing 2 economic policies. Higher stringency values indicate more economic support such as debt relief. Colour coding indicates the policy extent, with blue signalling more economic support, and red signalling less. Grey areas indicate states without data for this indicator.

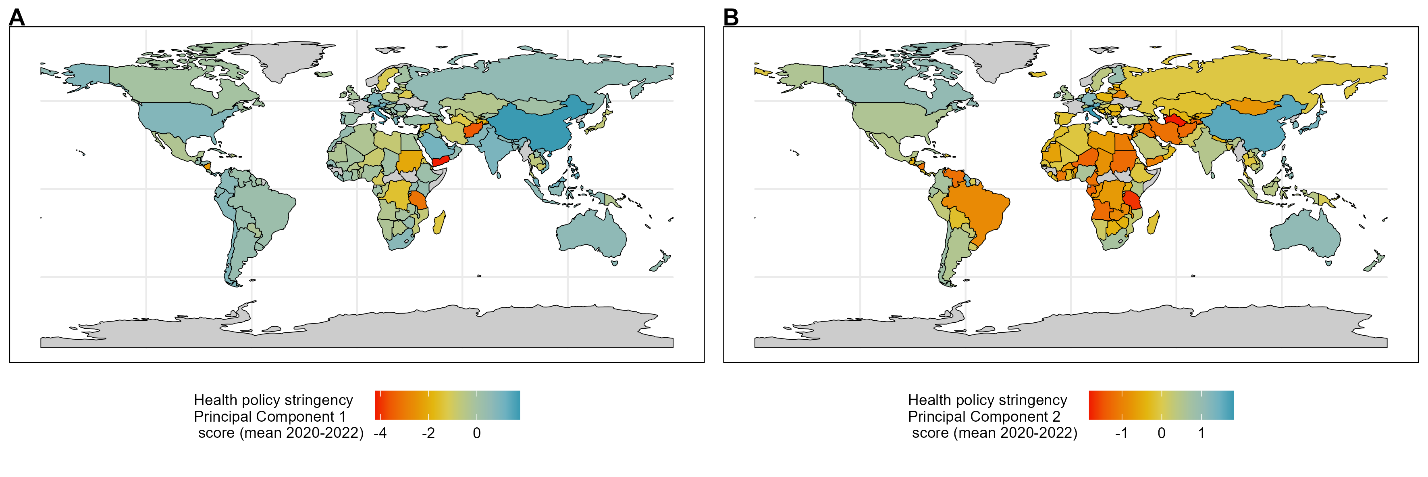

**Figure 10** Health policy stringency variation across countries, as quantified by two Principal Components (panel A and B) summarizing 8 health policies. Higher stringency values indicate more health policies, such as mask mandates. Colour coding indicates the policy extent, with red signalling stronger health policy restrictions, and blue signalling less. Grey areas indicate states without data for this indicator.

Health financing: government, external, and private sources

One may expect that increased financial investment results in the ability to fund more crucial healthcare resources – from facilities, to equipment, to health workforce, and service quality. Thus, greater health financing expenditure may be expected to have contributed to the development of improved health systems over time, and in turn, greater pandemic preparedness and systems maintenance during the pandemic.

The WHO quantify country-level health financing for health across three key sources of financing: government general health expenditure (codified as GGHE-D in WHO health financing datasets), domestic private health expenditure (codified as PVT-D; of which out-of-pocket expenditure is a key component), and external funding (codified as EXT; e.g., donors) [26]. Research has shown that the source of health financing can impact health outcomes [27], therefore we conduct analyses on the three core component sources rather than an aggregate.

To enable more relevant comparison across countries we use Purchasing Power Parities (PPPs) estimates of each health financing variable and calculate pre-pandemic expenditure (2015-2019 mean expenditure, excluding missing values). PPP converts different currencies and weights for costs of goods in a country to enable comparisons across countries. Estimates are also used per capita, to give a comparative level of investment across countries.

Additionally, we did explore the difference between pre-pandemic (2015-2019) and during pandemic (2020-2021; 2022 not available at time of analysis) and found minimal differences in trends. For consistency across other variables, and given hypothesis is tied to longer-term development of stronger health systems, we use pre-pandemic mean values only.

We visualise the underlying variation in country health financing from this dataset in **Figure 11** (government financing), **Figure 12** (private health financing) and **Figure 13** (external financing).

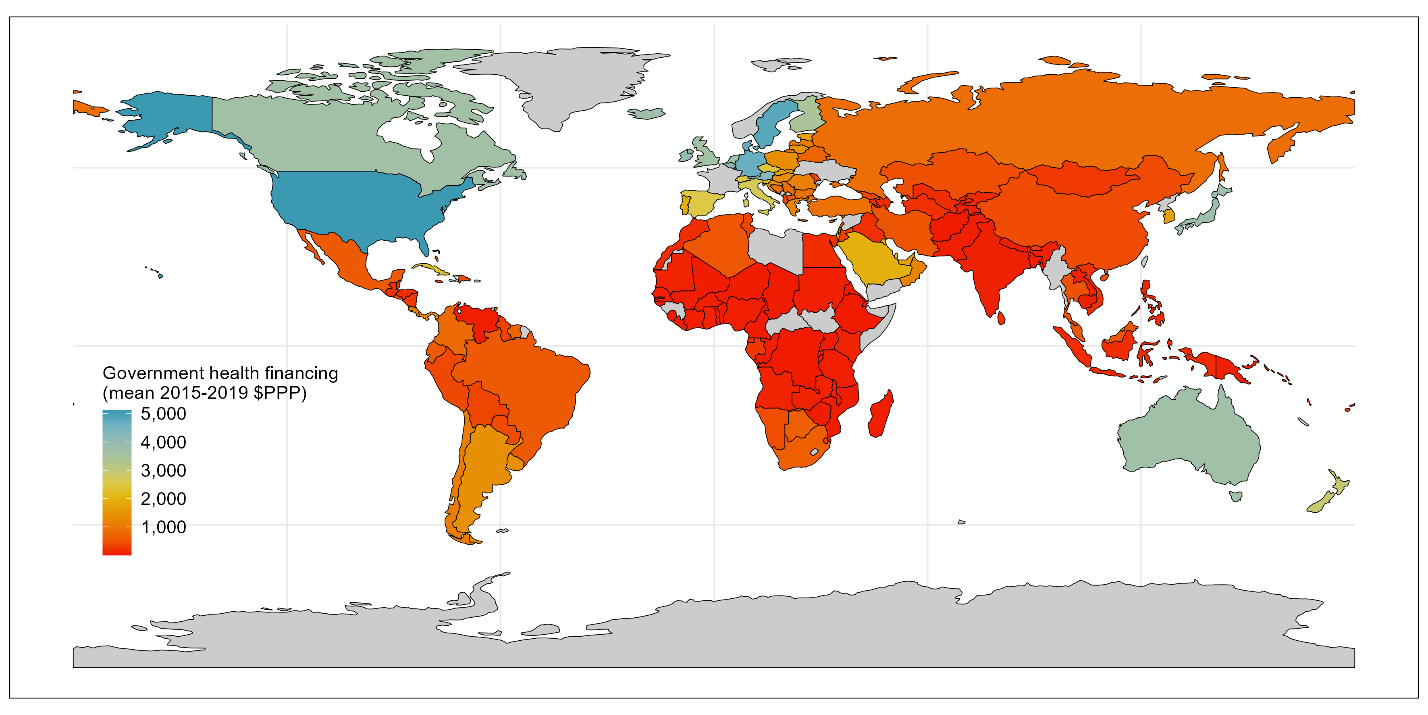

**Figure 11** Government health financing variation across countries, measured in terms of Purchasing Power Parity to enable comparison, from 2015-2019. Colour coding indicates the extent of government health financing, with blue signalling more investment, and red signalling less. Grey areas indicate states without data for this indicator.

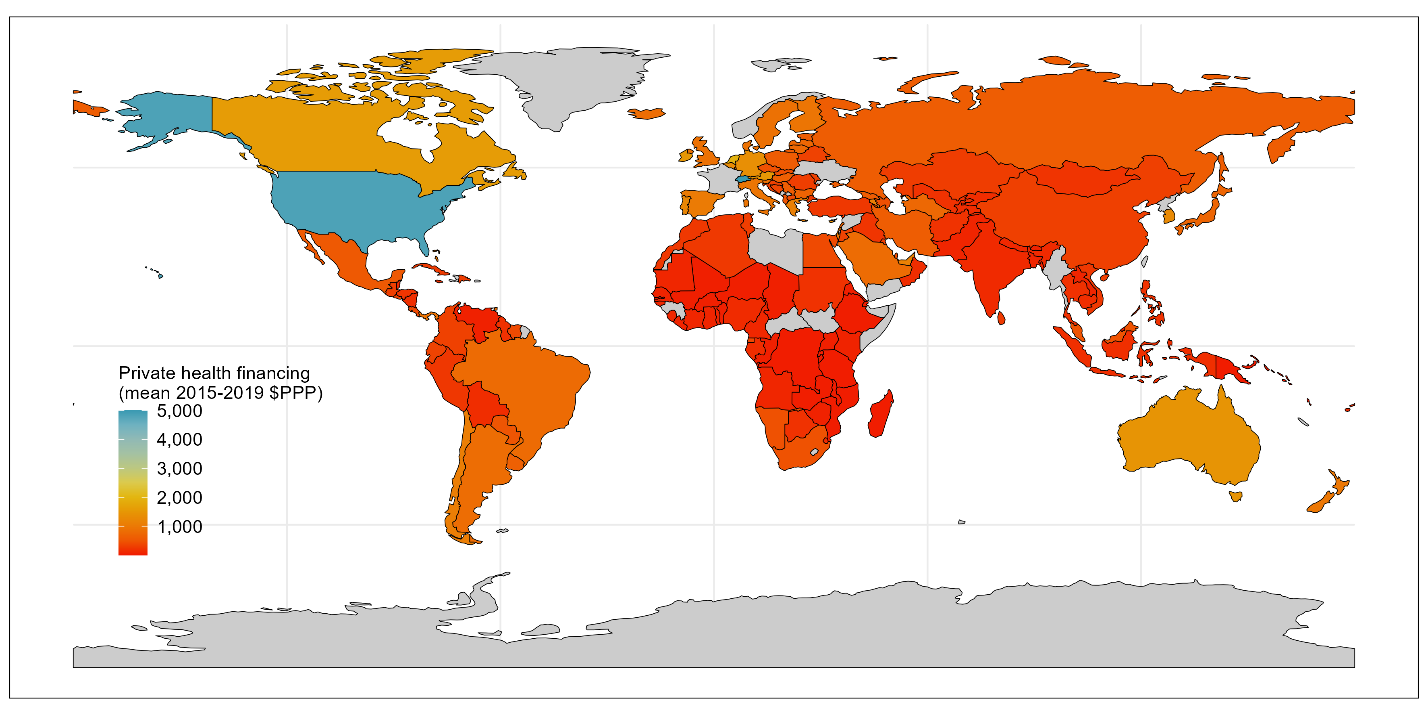

**Figure 12** Private health financing (e.g., out-of-pocket expenditure) variation across countries, measured in terms of Purchasing Power Parity to enable comparison, from 2015-2019. Colour coding indicates the extent of private health financing, with blue signalling more investment, and red signalling less. Grey areas indicate states without data for this indicator.

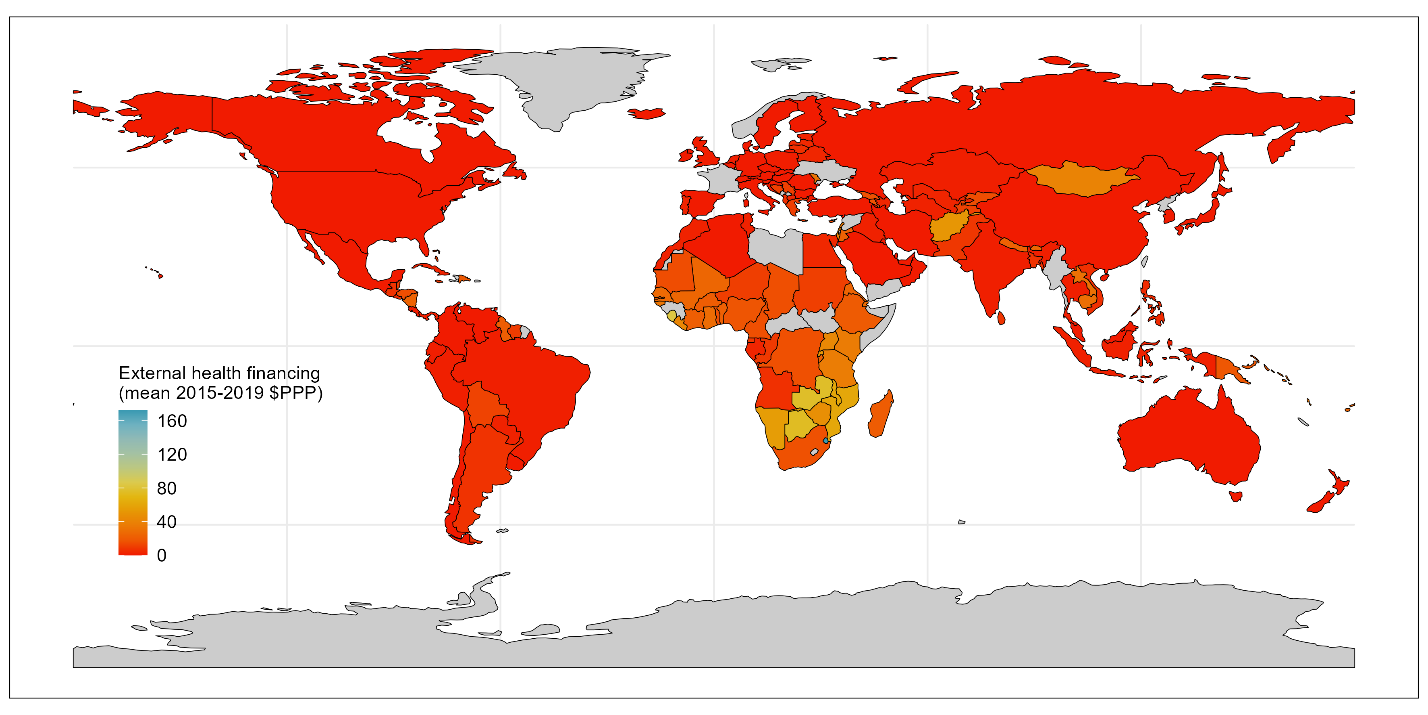

**Figure 13** External (e.g., donor) health financing variation across countries, measured in terms of Purchasing Power Parity to enable comparison, from 2015-2019. Colour coding indicates the extent of external health financing, with blue signalling more investment, and red signalling less. Grey areas indicate states without data for this indicator.

Mobility: Outside and residential

We hypothesis that regardless of cause of change of mobility – whether enforced through stay-at-home orders, or self-imposed through caution or fear of contracting COVID-19 through gatherings –countries exhibiting severely reduced movement outside the home during the pandemic, may have had or felt reduced access to health facilities (whether the facilities were in fact open or not).

To investigate this, we used mobile phone data from Google that tracked changes in movement to six different categories of places from 2020 to 2022 inclusive. Google report percent changes in movement compared to a baseline for the same day of the week per location for each day of the pandemic. We calculate the average annual change in mobility per location per year by taking the mean of the daily effect changes (excluding missing values). Google mobility data is aggregated, anonymised data collated from individuals with Google devices (typically Android mobile phones). As a result, the data may not be representative of all individuals in a given country or area, but does provide a large dataset and identification of broad movement trends, since it is compared to its own baseline from pre-pandemic.

We then investigated internal correlation amongst indicators and found that five of these categories were highly correlated to each other, namely the “outside” locations – retail and recreation, groceries and pharmacies, parks, transit stations, and workplaces. Inversely correlated to these five destinations was the “residential” indicator. For simplification of analysis, we produced a summary “outside” mobility indicator by conducting PCA on the five outside annualised indicators taking the mean of each of the five outdoor categories. The first PC described 81% of the variation in mobility change to outside locations and is comprised of a relatively even contribution (~20% each) of each underlying destination.

We visualise the underlying variation in country mobility from this dataset in **Figure 14** (outside) and **Figure 15** (residential).

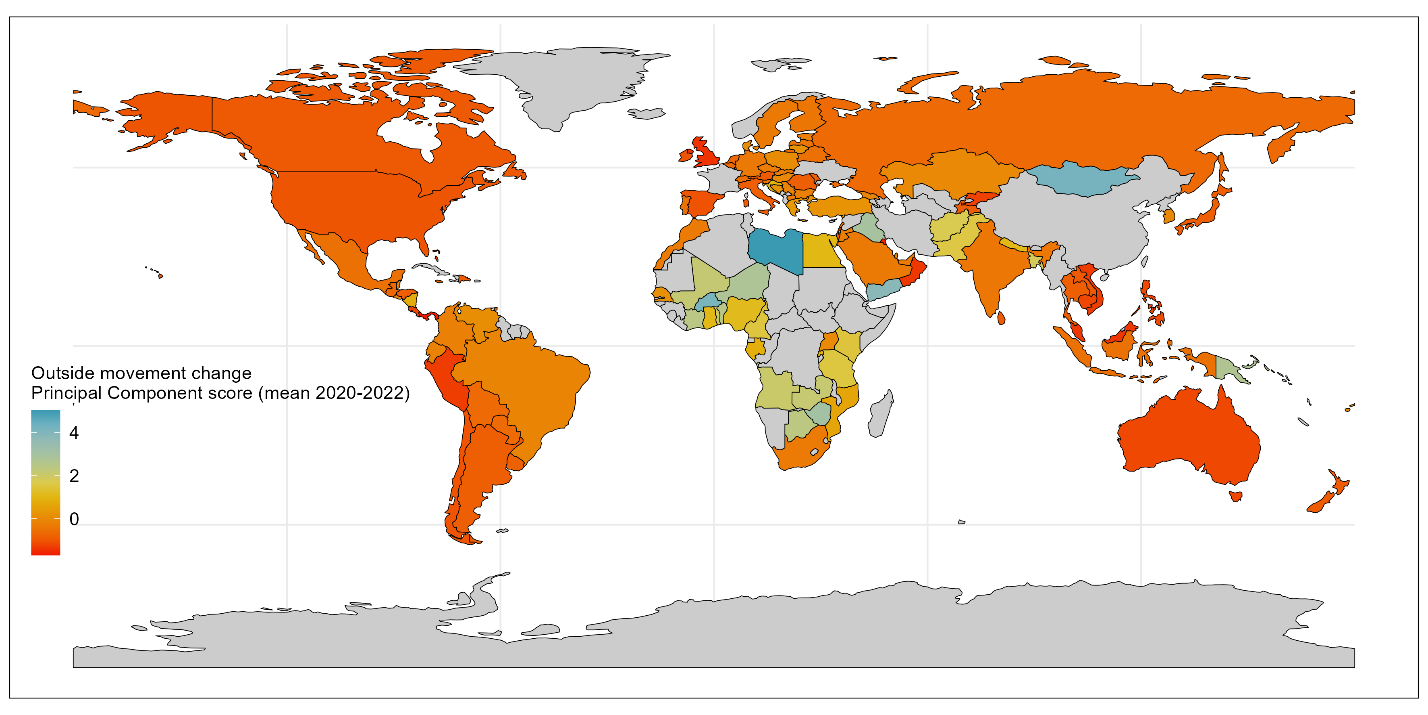

**Figure 14** Outside mobility change during pandemic variation across countries, as quantified by a single Principal Component summarizing movement to six locations (retail, grocery, parks, transit, and workplaces). Lower outside mobility values indicate greater reductions in outside movement during the pandemic. Higher stringency values indicate more health policies, such as mask mandates. Colour coding indicates the extent of outside mobility changes, with red signalling stronger declines, and blue signalling less. Grey areas indicate states without data for this indicator.

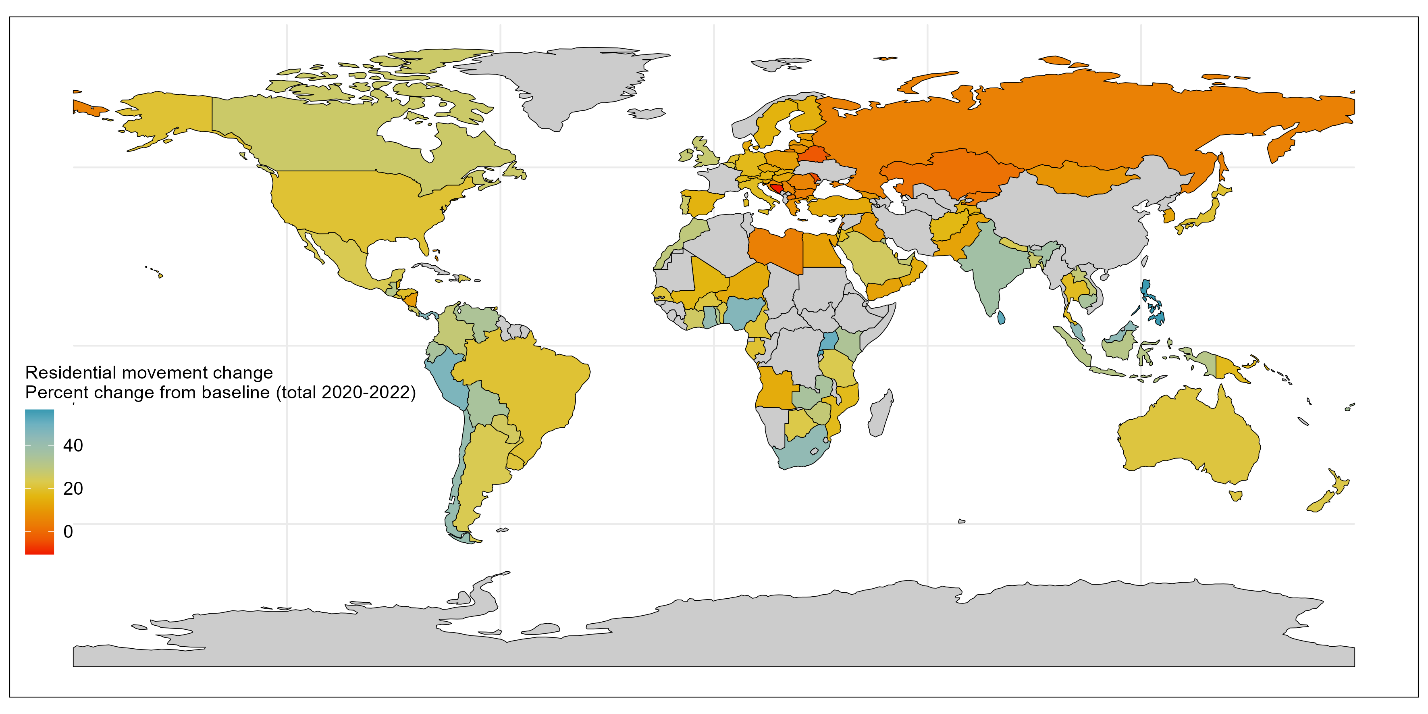

**Figure 15** Residential mobility change during pandemic variation across countries, as quantified by cumulative annual percentage change compared to baseline from Google phone data. Higher residential values (blue) indicate more stay-at-home behaviour during the pandemic and lower values (red) indicate less such behaviour, compared to baseline. Grey areas indicate states without data for this indicator.

Country wealth

Pandemic responses were cross-cutting and not specific to the health system, e.g., non-pharmaceutical interventions and border closures. As a result, it may be hypothesised that broader country access to financing may influence the ability to develop, maintain, and deploy effective pandemic responses. For this reason, we include Gross Domestic Product (GDP) as a predictor.

We take the GDP per capita scaled for PPP as an indicator of comparative country wealth, sourced from the World Bank [28]. As per the health financing indicators, this data is pre-quantified on a per capita scale and in terms of PPP. We use the 5-year mean GDP per capita from 2015-2019 (excluding missing values) to provide a baseline on comparative country wealth pre-pandemic, since GDP during the pandemic may reflect disruptive impact of the pandemic on the national economy.

We visualise the underlying variation in country wealth from this dataset in **Figure 16**.

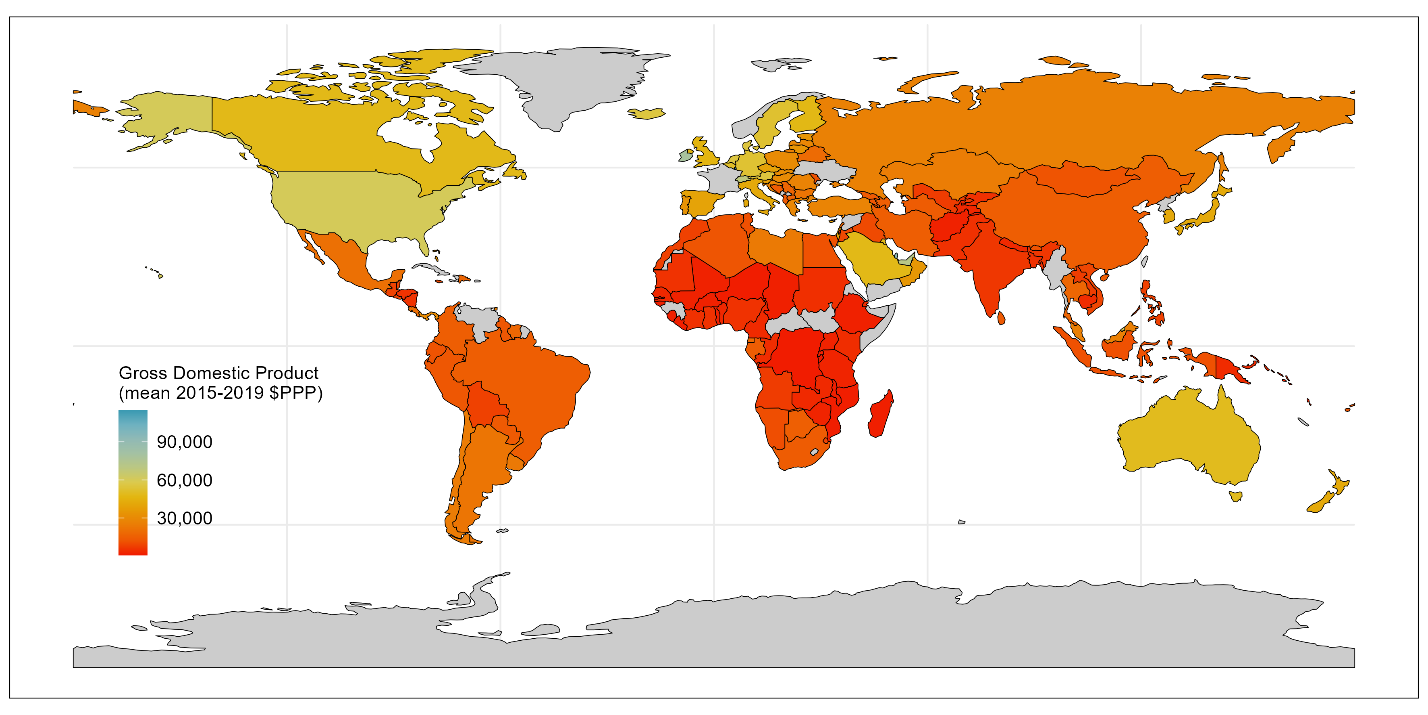

**Figure 16** Country wealth pre-pandemic variation across countries, as quantified by Gross Domestic Product (GDP) per capita in terms of Purchasing Power Parity (from 2015-2019 mean) from World Bank data. Higher GDP is coloured blue, and lower GDP are in red. Grey areas indicate states without data for this indicator.

C. **Univariate analyses**

Methodology

We systematically explore univariate relationships between each independent variable and the dependent variable. We visualise pairwise relationships, plotting three versions of each using Loess regression:

(1) **Per year**: Coverage deltas per year (y-axis) against each predictor (x-axis) plot either per year for pandemic varying factors (e.g., excess mortality which has mortality estimates for 2020, 2021, and 2022) or against the pre-pandemic average for variables quantified as pre-pandemic averages (e.g., GDP which is based on the mean GDP from 2015-2019 inclusive) per country. Three lines are fit, one for each year of the pandemic – to see if there is any variation in trends within the pandemic period.
(2) **Combined**: The same datapoints as first figure, with a single line of best fit across all years and all countries.
(3) **Total**: Cumulative coverage deltas summed over 3-years (y-axis) against each predictor (x-axis) plot either per year for pandemic varying factors or against the pre-pandemic average for variables quantified as pre-pandemic averages or PCs per country. Single line of best fit across all countries.

Univariate analyses are used to assess the existence of linear or non-linear relationships, and to verify if pairwise relationships varied over time or could be combined for multivariate analyses.

Overall results

Across most predictors, relationships per-year (e.g., **Figure 2 column A**) were similar to combined results (e.g., **Figure 2 column B**), allowing multivariate analyses to be conducted on all country-year datapoints together.

Pre-pandemic immunisation programme performance

Univariate analysis of pre-pandemic immunisation performance and coverage deltas during the pandemic indicated a non-linear relationship (**Figure 17**): below ~83% pre-pandemic DTP3 coverage, coverage deltas during the pandemic increase with increasing coverage (i.e., the inverse of our hypothesis); however, after ~83% coverage, higher pre-pandemic immunisation system performance is associated with reduced RI pandemic impact. This threshold effect suggests that there may be other explanatory variables that explain this intermediate effect, and/or that data quality and quantity in countries with the lowest reported coverage may be less robust and complicate interpretation.

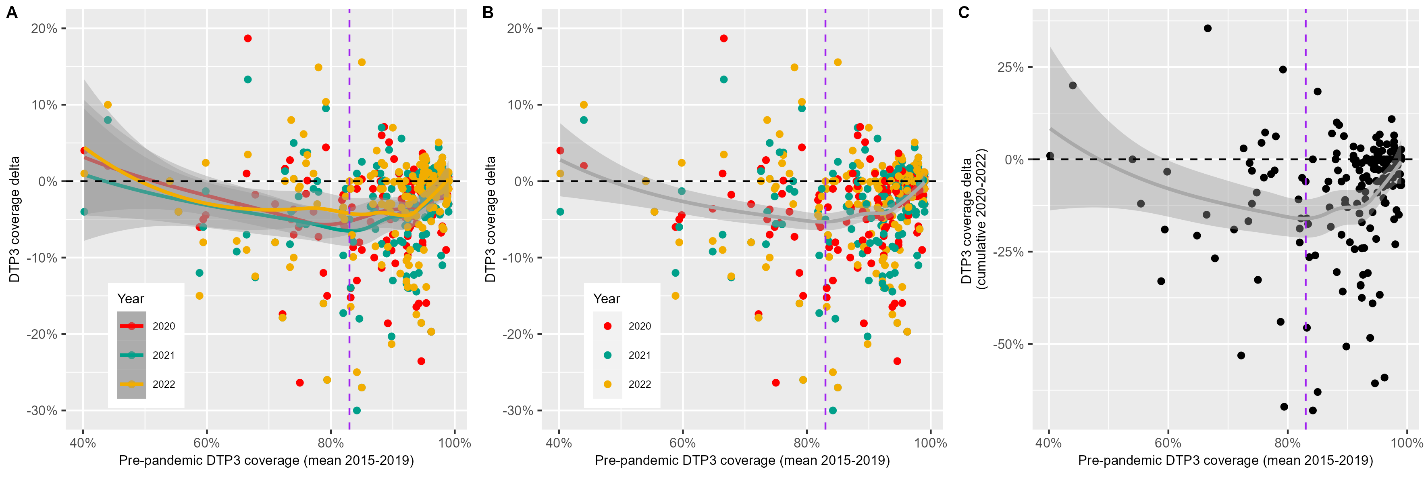

**Figure 17** Univariate analysis of coverage deltas (y-axis) against pre-pandemic immunisation programme performance, as quantified by mean DTP3 coverage from 2015-2019 (x-axis). Each dot represents a country-year datapoint, e.g., Senegal in 2021. For Panels A and B, dot colour indicates year of the pandemic – red for 2020, green for 2021, and yellow for 2022. Panel A (Per year) shows three lines of best fit for each year of the pandemic; Panel B (Combined) fits a single line of best fit for all years of data; and Panel C (Cumulative) plots total coverage deltas (the sum of each year). All fitted lines in the figure represent Loess (locally estimated scatterplot smoothing) curves, with the surrounding grey areas indicating the 95% confidence intervals for the fitted values. The horizontal black line indicates no change in DTP3 coverage. The vertical purple line indicates the visually-assessed threshold turning point.

Health workforce capacity

As per the pre-pandemic immunisation variable there is a threshold effect at 50 doctors and nurses per 10,000 population – see **Figure 18**. With pandemic immunisation performance decreasing until a threshold of 50 doctors and nurses is reached, and then performance increasing with increasing health workers.

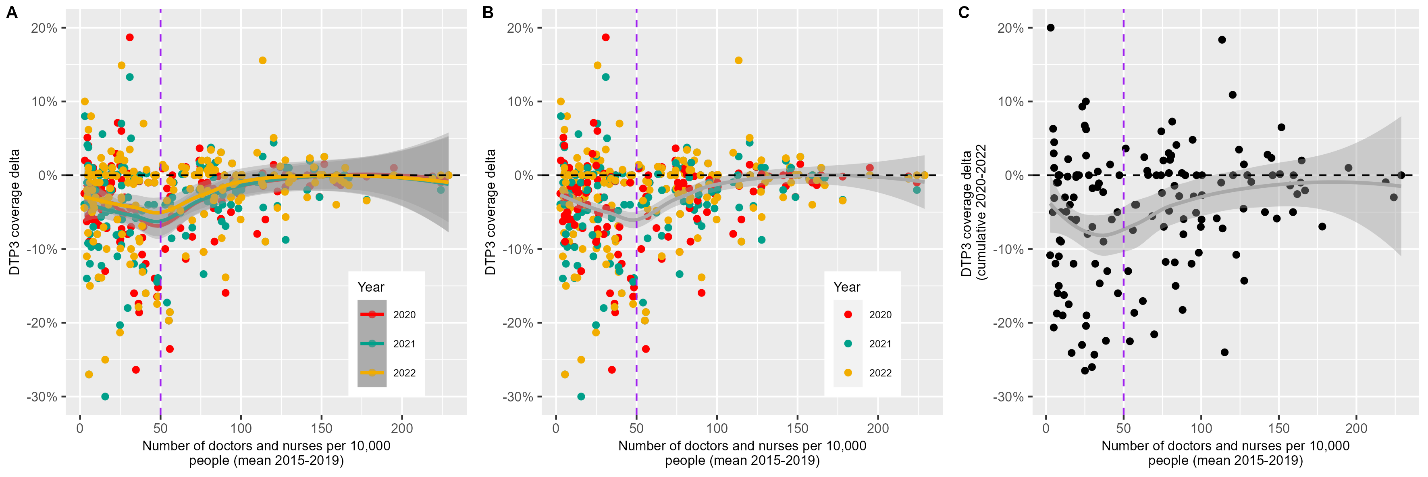

**Figure 18** Univariate analysis of coverage deltas (y-axis) against health workforce capacity, as quantified by mean number of doctors and nurses per 10,000 population from 2015-2019 (x-axis). Each dot represents a country-year datapoint, e.g., Senegal in 2021. For Panels A and B, dot colour indicates year of the pandemic – red for 2020, green for 2021, and yellow for 2022. Panel A (Per year) shows three lines of best fit for each year of the pandemic; Panel B (Combined) fits a single line of best fit for all years of data; and Panel C (Cumulative) plots total coverage deltas (the sum of each year). All fitted lines in the figure represent Loess (locally estimated scatterplot smoothing) curves, with the surrounding grey areas indicating the 95% confidence intervals for the fitted values. The horizontal black line indicates no change in DTP3 coverage. The vertical purple line indicates the visually-assessed threshold turning point.

Health systems strength

Univariate analysis of changes in immunisation coverage during the pandemic against the UHC index (**figure 19**) indicated a threshold affect at a UHC index of approximately 70. Before this point, DTP3 coverage delta is approximately flat, i.e., no change, with increasing UHC index. After a UHC index threshold score of 70, increases in the UHC index are associated with better pandemic routine immunisation maintenance.

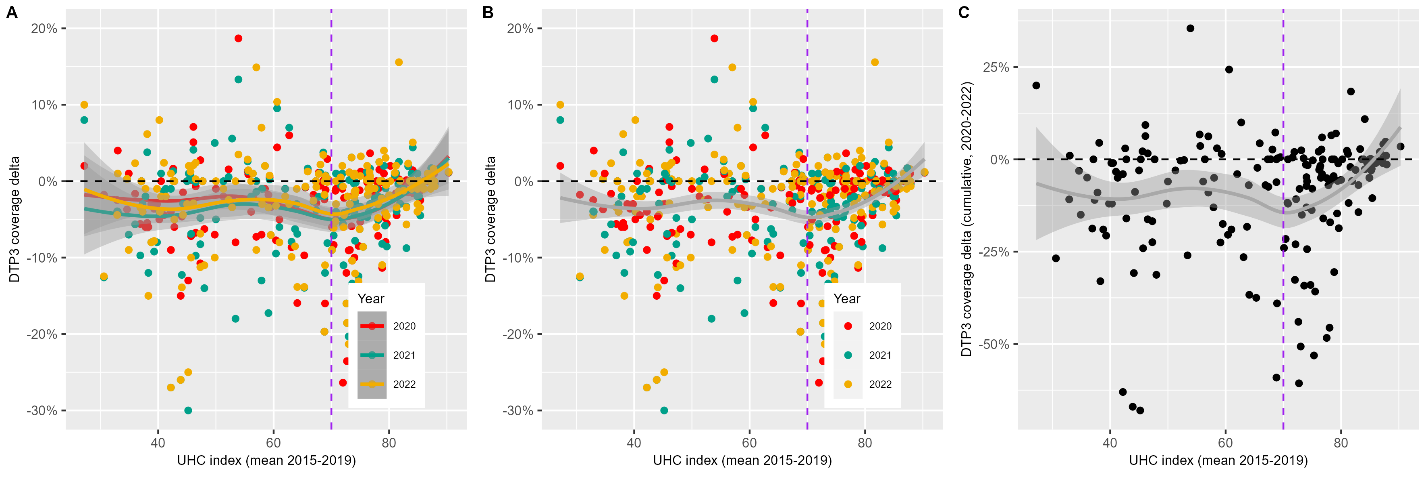

**Figure 19** Univariate analysis of coverage deltas (y-axis) against health systems strength, as quantified by the mean Universal Health Coverage index score from 2015-2019 (x-axis). Each dot represents a country-year datapoint, e.g., Senegal in 2021. For Panels A and B, dot colour indicates year of the pandemic – red for 2020, green for 2021, and yellow for 2022. Panel A (Per year) shows three lines of best fit for each year of the pandemic; Panel B (Combined) fits a single line of best fit for all years of data; and Panel C (Cumulative) plots total coverage deltas (the sum of each year). All fitted lines in the figure represent Loess (locally estimated scatterplot smoothing) curves, with the surrounding grey areas indicating the 95% confidence intervals for the fitted values. The horizontal black line indicates no change in DTP3 coverage. The vertical purple line indicates the visually-assessed threshold turning point.

Global Health Security

Univariate analysis (**Figure 20**) indicated a less clear relationship between DTP3 coverage deltas and GHSI score, with a potential non-linear relationship visible when assessed cumulatively rather than per year. Cumulatively, a threshold effect was seen such that pandemic coverage performance decreased until a GHSI score of ~35 and then improved with increasing GHSI score.

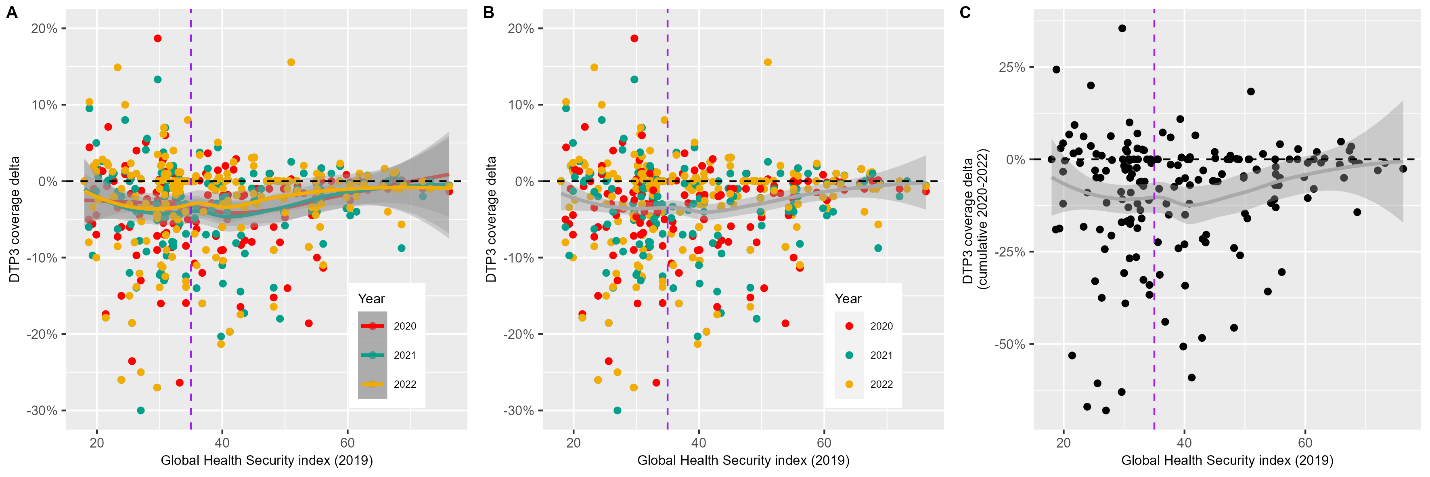

**Figure 20** Univariate analysis of coverage deltas (y-axis) against Global Health Security strength, as quantified by the Global Health Security index score (x-axis). Each dot represents a country-year datapoint, e.g., Senegal in 2021. For Panels A and B, dot colour indicates year of the pandemic – red for 2020, green for 2021, and yellow for 2022. Panel A (Per year) shows three lines of best fit for each year of the pandemic; Panel B (Combined) fits a single line of best fit for all years of data; and Panel C (Cumulative) plots total coverage deltas (the sum of each year). All fitted lines in the figure represent Loess (locally estimated scatterplot smoothing) curves, with the surrounding grey areas indicating the 95% confidence intervals for the fitted values. The horizontal black line indicates no change in DTP3 coverage. The vertical purple line indicates the visually-assessed threshold turning point.

COVID-19 vaccination

On an annual basis, univariate analysis indicated a roughly linear relationship between COVID-19 vaccination and DTP3 coverage deltas (**Figure 21, panel B**), such that the greater the number of COVID-19 vaccines administered per capita, the stronger the maintenance of RI coverage during the pandemic. When evaluated cumulatively across all 3-years of the pandemic, a non-linear relationship was observed, with pandemic RI performance declining until a threshold of approximately 1 COVID-19 vaccination per person, and then improvements thereafter (**Figure 21, panel C**).

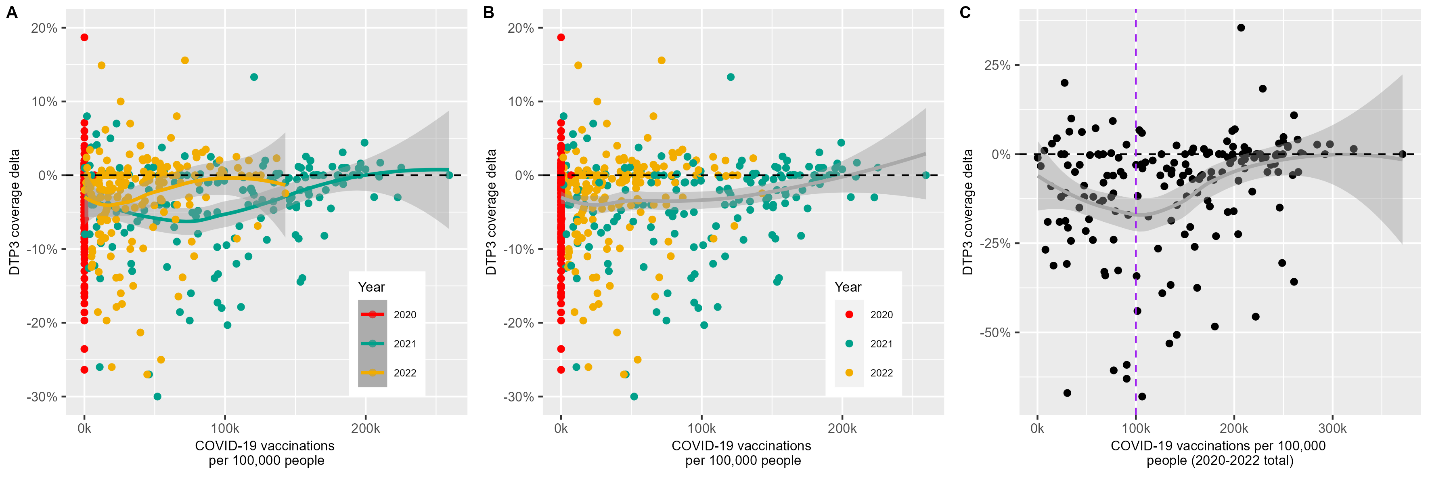

**Figure 21** Univariate analysis of coverage deltas (y-axis) against COVID-19 vaccination levels, as quantified by number of COVID-19 vaccines administered per 100,000 population (x-axis). Each dot represents a country-year datapoint, e.g., Senegal in 2021. For Panels A and B, dot colour indicates year of the pandemic – red for 2020, green for 2021, and yellow for 2022. Panel A (Per year) shows three lines of best fit for each year of the pandemic; Panel B (Combined) fits a single line of best fit for all years of data; and Panel C (Cumulative) plots total coverage deltas (the sum of each year). All fitted lines in the figure represent Loess (locally estimated scatterplot smoothing) curves, with the surrounding grey areas indicating the 95% confidence intervals for the fitted values. The horizontal black line indicates no change in DTP3 coverage. The vertical purple line indicates the visually-assessed threshold turning point.

COVID-19 disease (excess mortality)

Univariate analysis indicated a non-linear relationship between excess mortality during the pandemic and DTP3 coverage deltas. This relationship was more pronounced when looking at cumulative trends (**Figure 22, Panel C**). Below approximately 200 excess deaths per 100,000 people per year DTP3 coverage deltas increase with increased excess mortality, and then decrease until around 300 excess deaths per 100,000 people. After this threshold, the relationship is relatively flat with minimal change in DTP3 coverage with further increases in excess mortality.

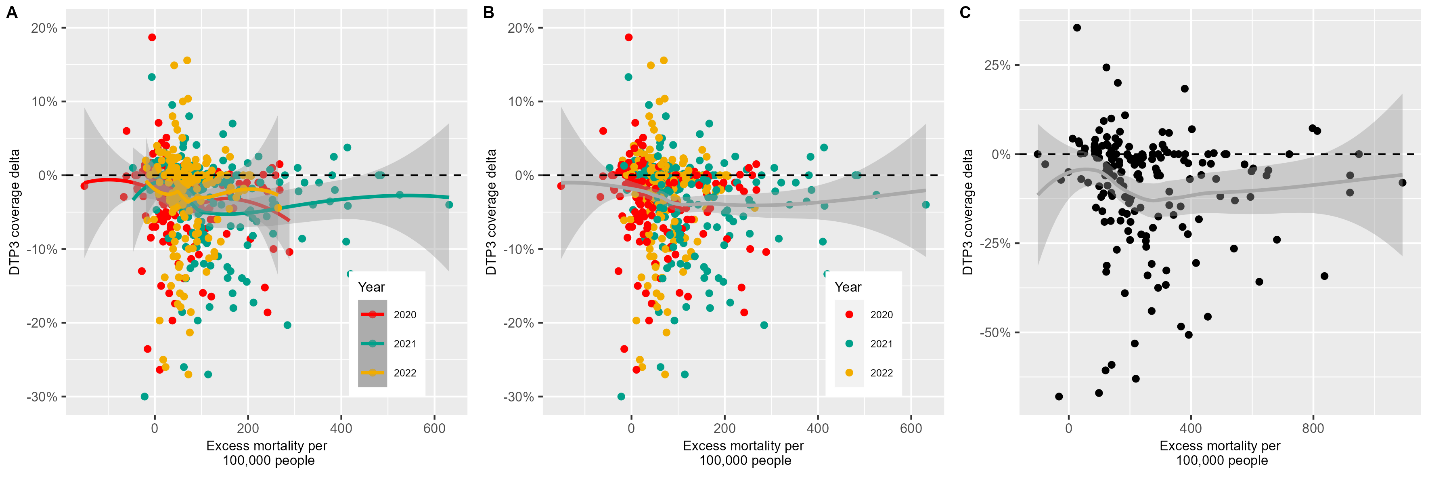

**Figure 22** Univariate analysis of coverage deltas (y-axis) against COVID-19 disease, as quantified by excess mortality per 100,000 people (x-axis). Each dot represents a country-year datapoint, e.g., Senegal in 2021. For Panels A and B, dot colour indicates year of the pandemic – red for 2020, green for 2021, and yellow for 2022. Panel A (Per year) shows three lines of best fit for each year of the pandemic; Panel B (Combined) fits a single line of best fit for all years of data; and Panel C (Cumulative) plots total coverage deltas (the sum of each year). All fitted lines in the figure represent Loess (locally estimated scatterplot smoothing) curves, with the surrounding grey areas indicating the 95% confidence intervals for the fitted values. The horizontal black line indicates no change in DTP3 coverage.

Pandemic policy: containment

The relationship between the containment policy PC and DTP3 coverage deltas is unclear (**Figure 23**), with a relatively flat relationship except at either extremes (very stringent or minimal policies) where DTP3 coverage deltas appear to increase with the most stringent or the weakest policies.

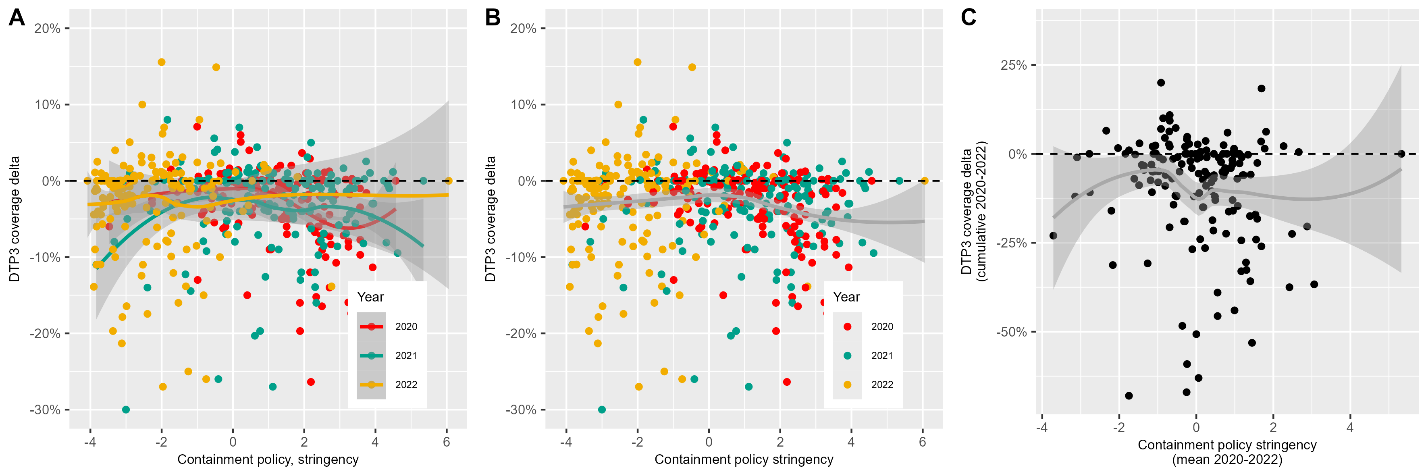

**Figure 23** Univariate analysis of coverage deltas (y-axis) against containment policy stringency, as quantified by a single Principal Component summarising eight containment policies (x-axis). Higher stringency values indicate more stringent containment policies such as more frequent or stringent school or workplace closures. Each dot represents a country-year datapoint, e.g., Senegal in 2021. For panels A and B, dot colour indicates year of the pandemic – red for 2020, green for 2021, and yellow for 2022. Panel A (Per year) shows three lines of best fit for each year of the pandemic; Panel B (Combined) fits a single line of best fit for all years of data; and Panel C (Cumulative) plots total coverage deltas (the sum of each year). All fitted lines in the figure represent Loess (locally estimated scatterplot smoothing) curves, with the surrounding grey areas indicating the 95% confidence intervals for the fitted values. The horizontal black line indicates no change in DTP3 coverage.

Pandemic policy: economic

There is no evidence of a relationship between the economic index PC and DTP3 coverage deltas (**Figure 24**) – the line of best fit across all versions of the plots are essentially horizontal.

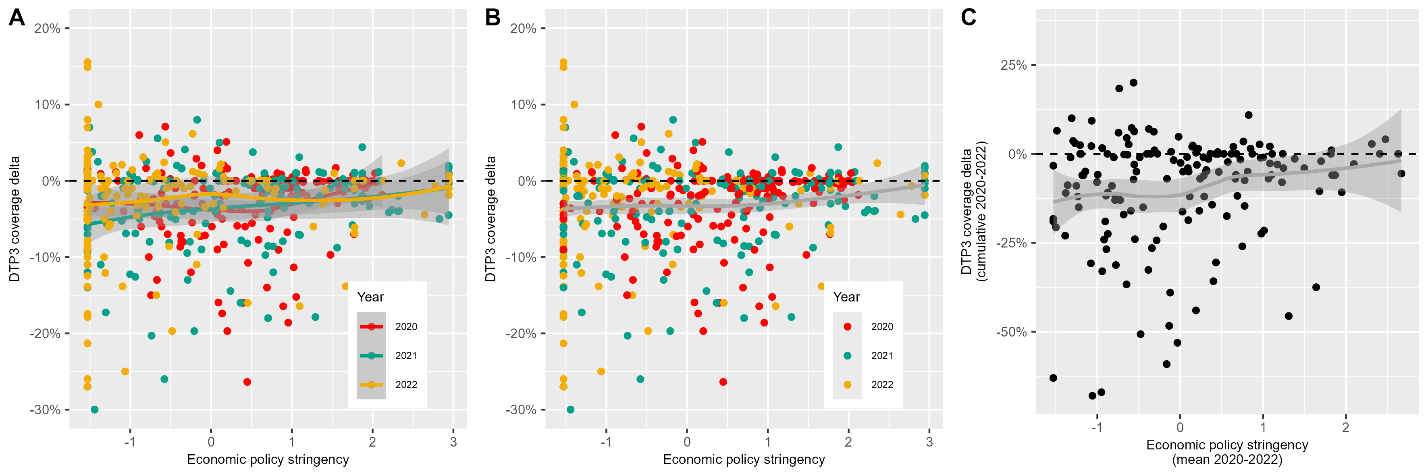

**Figure 24** Univariate analysis of coverage deltas (y-axis) against economic policy stringency, as quantified by a single Principal Component summarising two economic policies (x-axis). Higher stringency values indicate greater economic support e.g., broader economic relief during the pandemic. Each dot represents a country-year datapoint, e.g., Senegal in 2021. For Panels A and B, dot colour indicates year of the pandemic – red for 2020, green for 2021, and yellow for 2022. Panel A (Per year) shows three lines of best fit for each year of the pandemic; Panel B (Combined) fits a single line of best fit for all years of data; and Panel C (Cumulative) plots total coverage deltas (the sum of each year). All fitted lines in the figure represent Loess (locally estimated scatterplot smoothing) curves, with the surrounding grey areas indicating the 95% confidence intervals for the fitted values. The horizontal black line indicates no change in DTP3 coverage.

Pandemic policy: health

Univariate analysis indicated the lack of relationship between both health policy principal components and DTP3 coverage deltas (**Figure 25**), with minimal variation in DTP3 coverage as health policy stringency scores change.

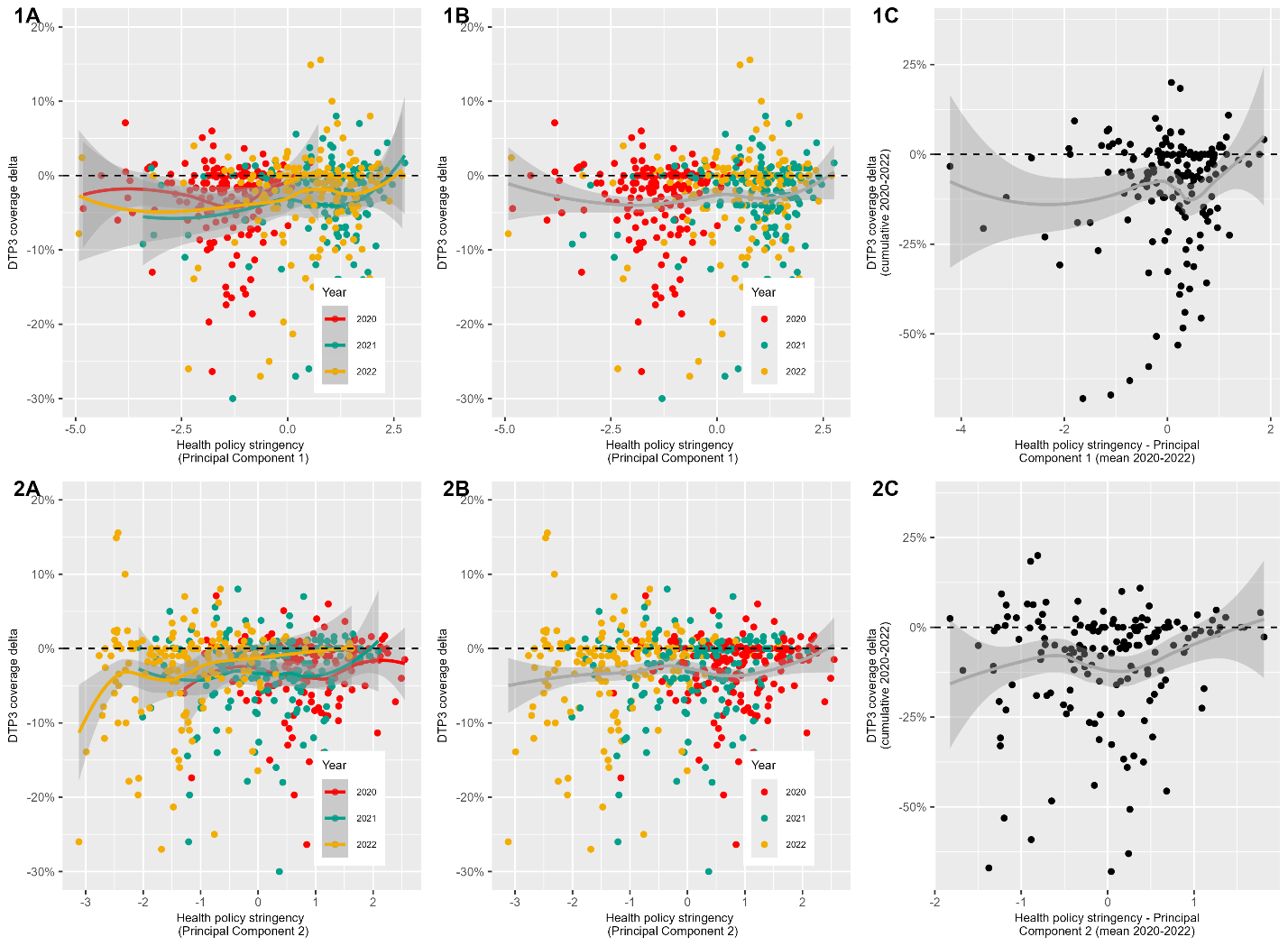

**Figure 25** Univariate analysis of coverage deltas (y-axis) against pandemic health policy stringency, as quantified by two Principal Components (PCs) summarising six health policies (x-axis: row 1 shows PC1 and row 2 shows PC2 ). Higher PC values indicate more stringent health policies such as requirements to wear masks more often. Each dot represents a country-year datapoint, e.g., Senegal in 2021. For Panels A and B, dot colour indicates year of the pandemic – red for 2020, green for 2021, and yellow for 2022. Panel A (Per year) shows three lines of best fit for each year of the pandemic; Panel B (Combined) fits a single line of best fit for all years of data; and Panel C (Cumulative) plots total coverage deltas (the sum of each year). All fitted lines in the figure represent Loess (locally estimated scatterplot smoothing) curves, with the surrounding grey areas indicating the 95% confidence intervals for the fitted values. The horizontal black line indicates no change in DTP3 coverage.

Mobility: outside

A strong positive relationship between outside mobility and DTP3 coverage deltas is seen below the PC value of approximately -1.5 (**Figure 26**). Outside mobility is encoded such that more negative scores indicate the greatest reductions in movement to outside locations (parks, transit, workplace, recreation, retail) thus for countries with strong reductions in movement to these locations, a greater decline in DTP3 coverage was observed. After this turning point the relationship plateaus.

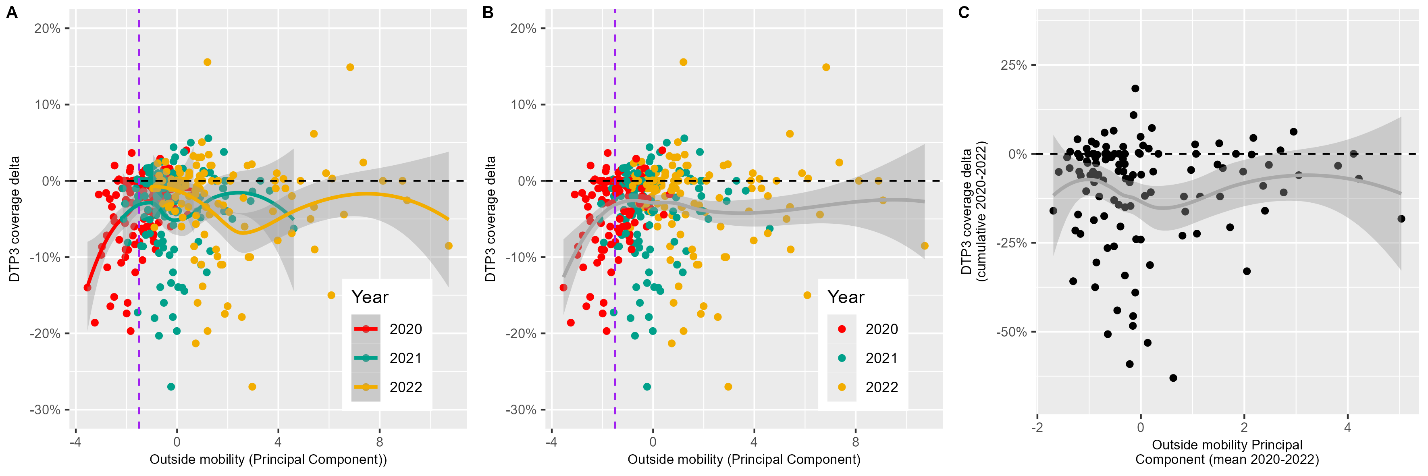

**Figure 26** Univariate analysis of coverage deltas (y-axis) against outside mobility, as quantified by a single Principal Component summarising change in movement to five locations from Google phone data (x-axis). Lower (more negative) outside mobility values indicate greater reductions in outside movement during the pandemic. Each dot represents a country-year datapoint, e.g., Senegal in 2021. Dot colour indicates year of the pandemic – red for 2020, green for 2021, and yellow for 2022. Panel A (Per year) shows three lines of best fit for each year of the pandemic; and Panel B (Combined) fits a single line of best fit for all years of data. All fitted lines in the figure represent Loess (locally estimated scatterplot smoothing) curves, with the surrounding grey areas indicating the 95% confidence intervals for the fitted values. The horizontal black line indicates no change in DTP3 coverage. The vertical purple line indicates the visually-assessed turning point.

Mobility: residential

Limited relationship between residential mobility and DTP3 coverage deltas is observed across each of the years – the line of best fit is relatively horizontal, see **Figure 27**.

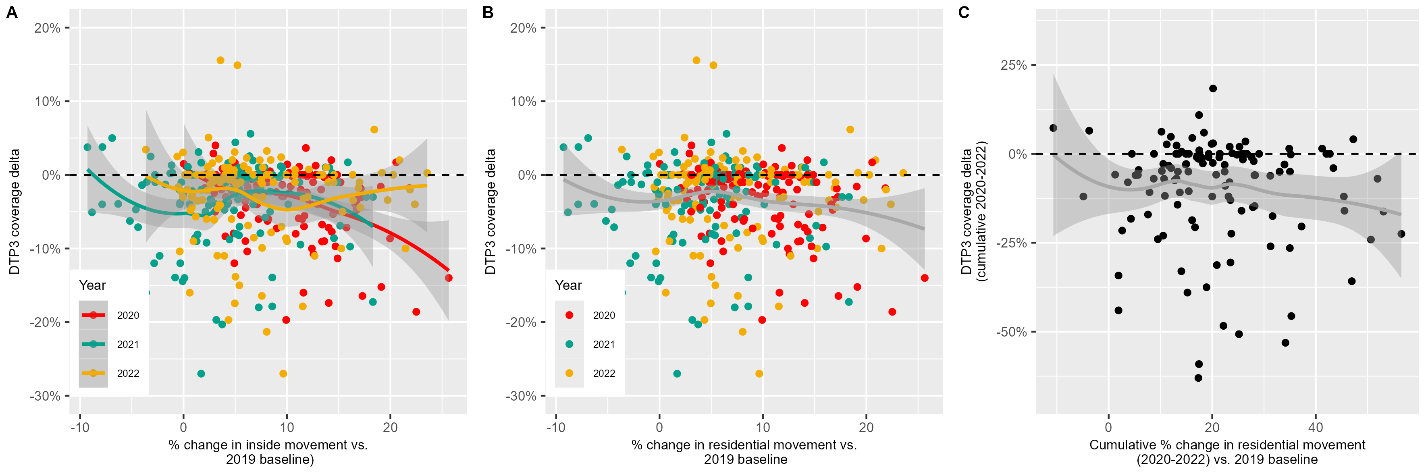

**Figure 27** Univariate analysis of coverage deltas (y-axis) against residential mobility, as quantified change in movement compared to pre-pandemic from Google phone data (x-axis). Higher percentage changes indicate more staying at home. Each dot represents a country-year datapoint, e.g., Senegal in 2021. Dot colour indicates year of the pandemic – red for 2020, green for 2021, and yellow for 2022. Panel A (Per year) shows three lines of best fit for each year of the pandemic; and Panel B (Combined) fits a single line of best fit for all years of data. All fitted lines in the figure represent Loess (locally estimated scatterplot smoothing) curves, with the surrounding grey areas indicating the 95% confidence intervals for the fitted values. The horizontal black line indicates no change in DTP3 coverage. The vertical purple line indicates the visually-assessed turning point.

Health financing: government

The relationship between government health financing and DTP3 coverage deltas is difficult to see across all data points – see **Figure 28 Panels A-C**, since outliers (with large investment) appear to contribute greatly to line fitting. Once zoomed in (**Panel D**) there appears to be some linear relationship such that after around $450 PPP of government health financing until around $2000 PPP pandemic immunisation performance improves with increasing investment. Below $450 PPP investment the trend is either minimal or worsening, and after $2000 PPP, the relationship plateaus with further increases in government health expenditure being associated with minimal changes in DTP3 coverage deltas.

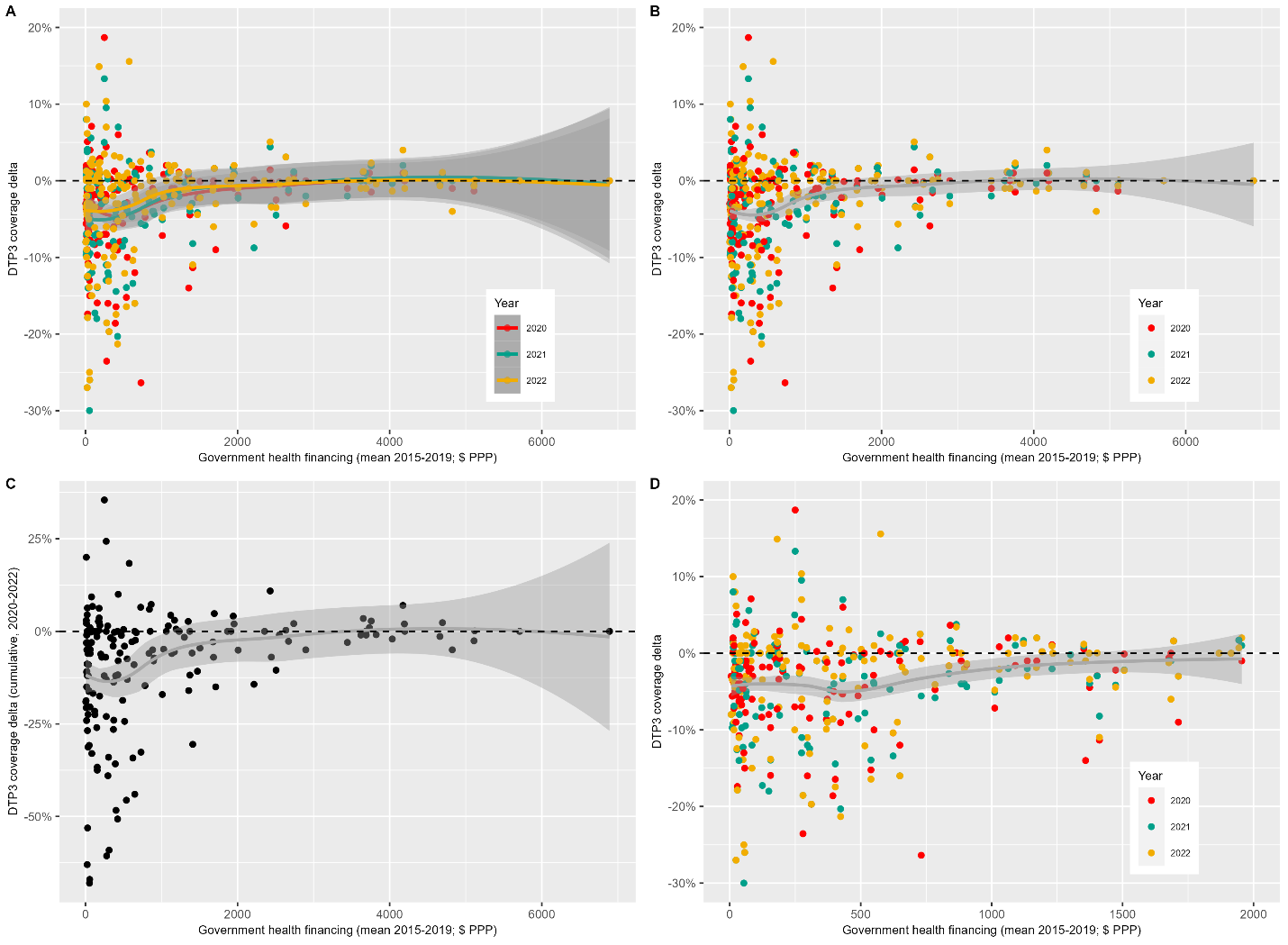

**Figure 28** Univariate analysis of coverage deltas (y-axis) against government health financing, as quantified by mean government health expenditure 2015-2019 weighted by Purchasing Power Parity per capita (x-axis). Each dot represents a country-year datapoint, e.g., Senegal in 2021. For Panels A, B, and D dot colour indicates year of the pandemic – red for 2020, green for 2021, and yellow for 2022. Panel A (Per year) shows three lines of best fit for each year of the pandemic; Panel B (Combined) fits a single line of best fit for all years of data; Panel C (Cumulative) plots total coverage deltas (the sum of each year); and Panel D is a zoomed in version of Panel B given the long tail in health financing range to allow easier identificaiton of trends. All fitted lines in the figure represent Loess (locally estimated scatterplot smoothing) curves, with the surrounding grey areas indicating the 95% confidence intervals for the fitted values. The horizontal black line indicates no change in DTP3 coverage. The vertical purple line indicates the visually-assessed threshold turning point.

Health financing: private

The relationship between private health financing expenditure and DTP3 coverage deltas is difficult to see across all data points, since outliers (with large investment) appear to contribute greatly to line fitting – **Figure 29 Panels A-C**. Across all data points there appears to be a parabolic relationship below $1000 PPP expenditure, with a turning point around $450 PPP. However, once zoomed in (**Panel D**) the relationship appears more flatlined below $450 PPP investment and from $450 to $1500 PPP private health expenditure it appears that pandemic immunisation performance improves with increasing private health care spend.

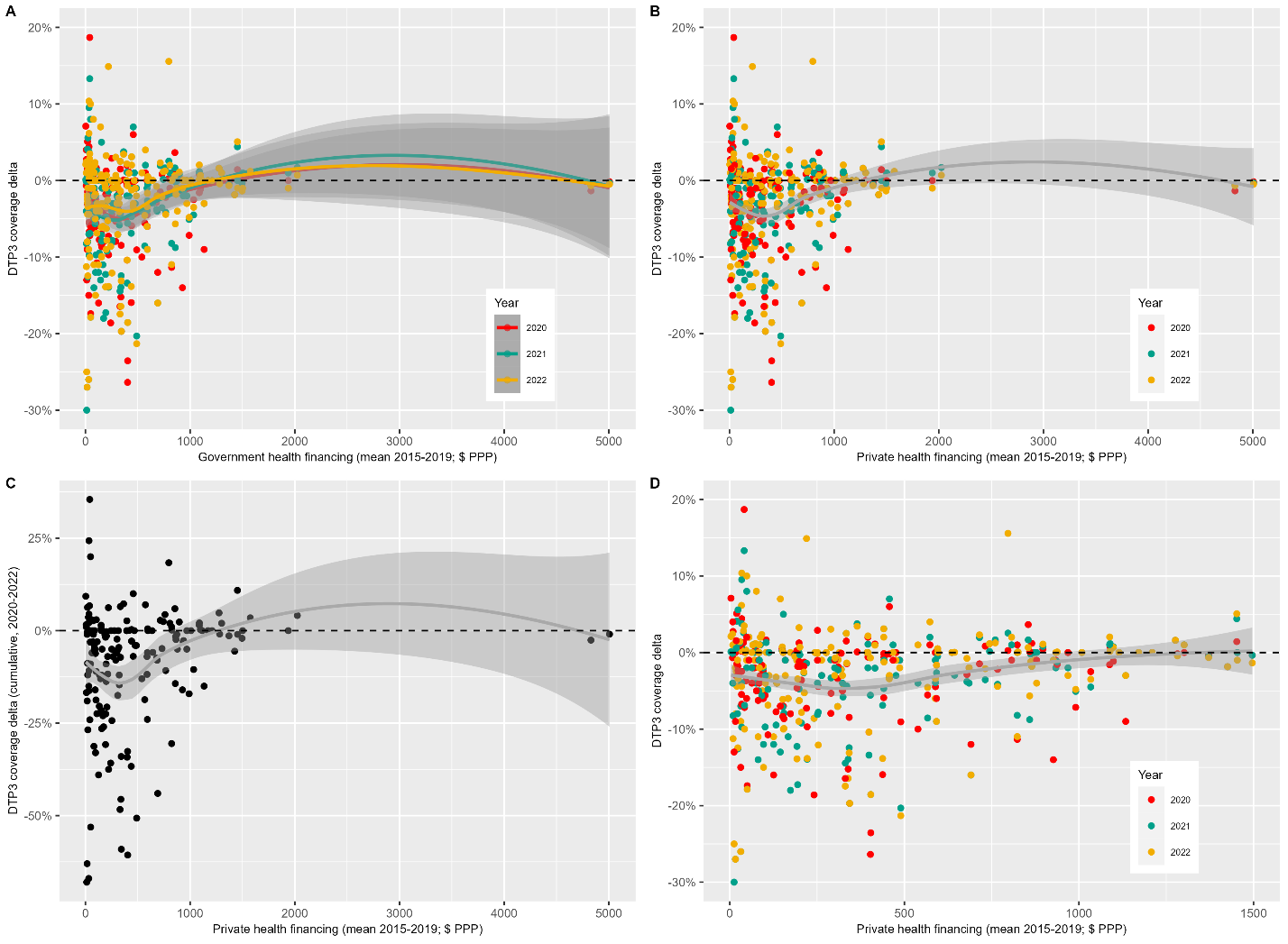

**Figure 29** Univariate analysis of coverage deltas (y-axis) against private health financing, as quantified by mean private health expenditure 2015-2019 weighted by Purchasing Power Parity per capita (x-axis). Each dot represents a country-year datapoint, e.g., Senegal in 2021. For Panels A, B, and D dot colour indicates year of the pandemic – red for 2020, green for 2021, and yellow for 2022. Panel A (Per year) shows three lines of best fit for each year of the pandemic; Panel B (Combined) fits a single line of best fit for all years of data; Panel C (Cumulative) plots total coverage deltas (the sum of each year); and Panel D is a zoomed in version of Panel B given the long tail in health financing range to allow easier identificaiton of trends. All fitted lines in the figure represent Loess (locally estimated scatterplot smoothing) curves, with the surrounding grey areas indicating the 95% confidence intervals for the fitted values. The horizontal black line indicates no change in DTP3 coverage. The vertical purple line indicates the visually-assessed threshold turning point.

Health financing: external

Once again, the relationship between external health financing expenditure and DTP3 coverage deltas is difficult to see across all data points, since outliers (with large investment) appear to contribute greatly to line fitting – **Figure 30 Panels A-C**. Once zoomed in (**Panel D**) the relationship appears relatively horizontal, with minimal trend apparent.

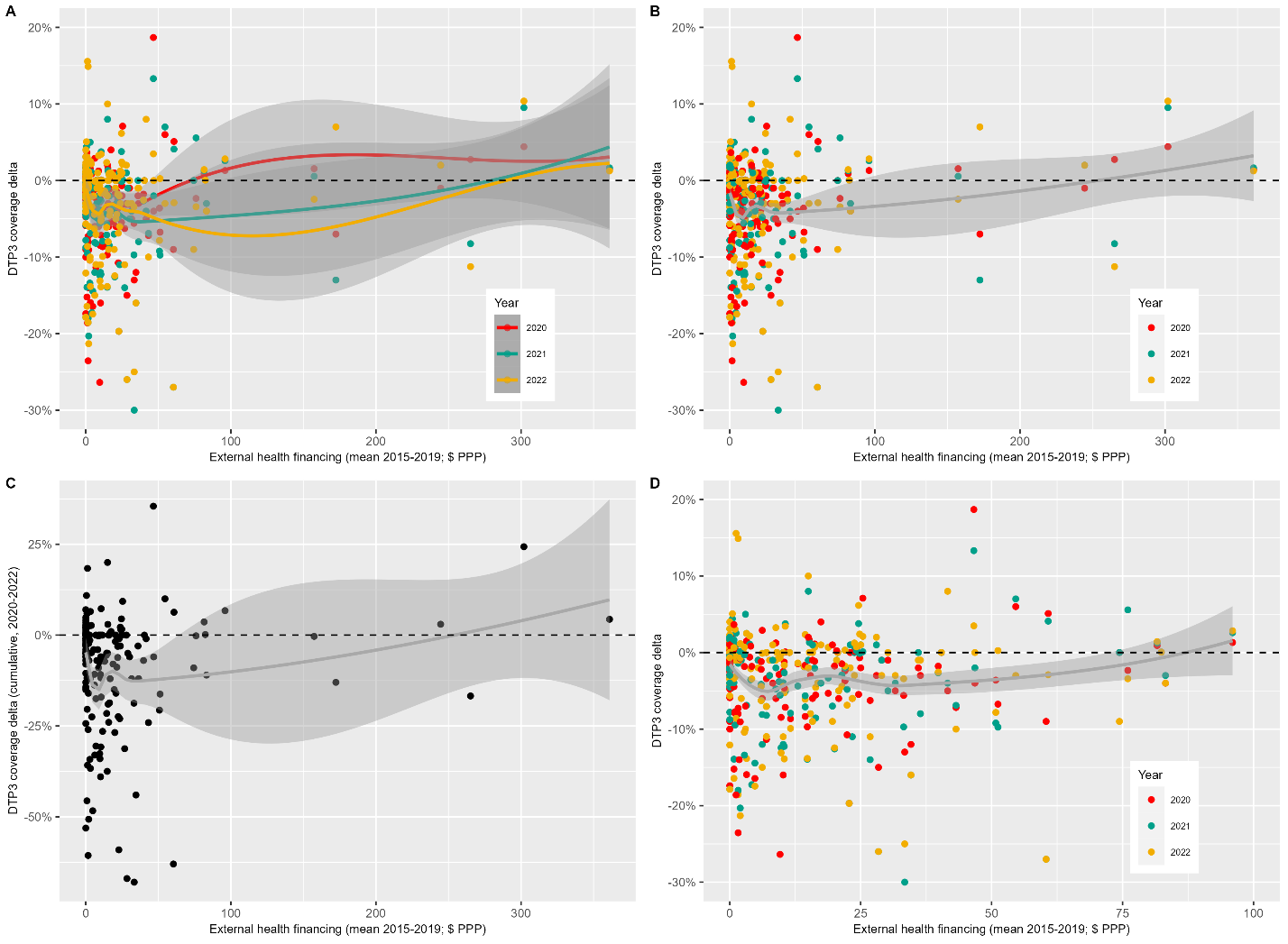

**Figure 30** Univariate analysis of coverage deltas (y-axis) against external (e.g., donor) health financing, as quantified by mean external health expenditure 2015-2019 weighted by Purchasing Power Parity per capita (x-axis). Each dot represents a country-year datapoint, e.g., Senegal in 2021. For Panels A, B, and D dot colour indicates year of the pandemic – red for 2020, green for 2021, and yellow for 2022. Panel A (Per year) shows three lines of best fit for each year of the pandemic; Panel B (Combined) fits a single line of best fit for all years of data; Panel C (Cumulative) plots total coverage deltas (the sum of each year); and Panel D is a zoomed in version of Panel B given the long tail in health financing range to allow easier identificaiton of trends. All fitted lines in the figure represent Loess (locally estimated scatterplot smoothing) curves, with the surrounding grey areas indicating the 95% confidence intervals for the fitted values. The horizontal black line indicates no change in DTP3 coverage. The vertical purple line indicates the visually-assessed threshold turning point.

Country wealth

Univariate analysis shows a roughly linear relationship between GDP and DTP3 coverage performance after approximately 15,000 $ PPP GDP per capita with increased GDP being associated with better vaccination maintenance during the pandemic – **Figure 31**. Below this threshold RI performance either slightly worsens with increasing GDP or appears relatively flat.

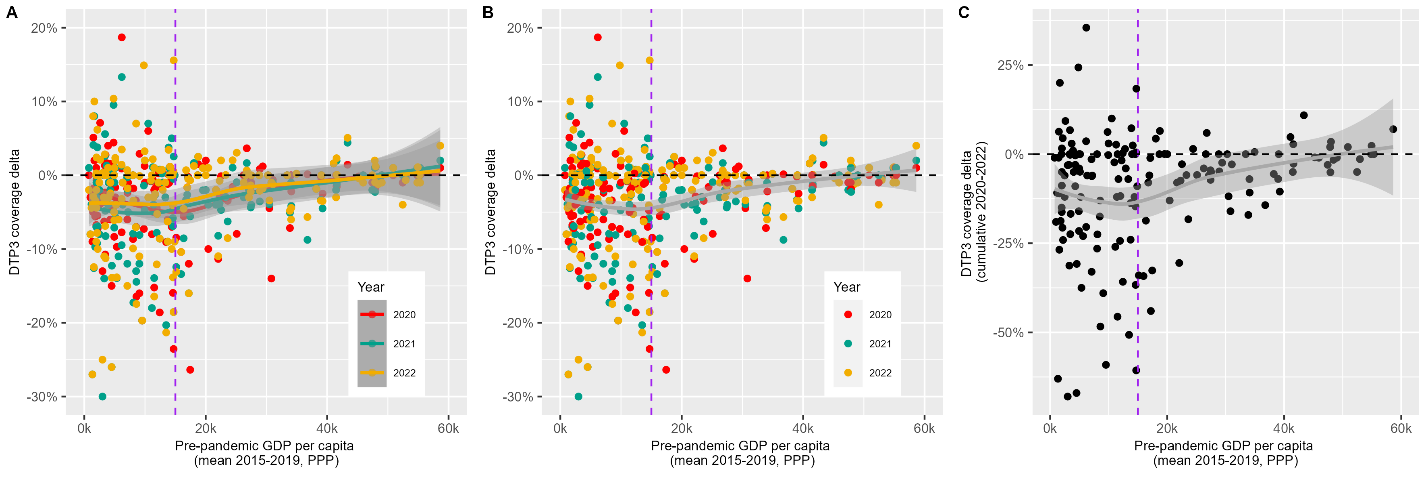

**Figure 31** Univariate analysis of coverage deltas (y-axis) against country wealth, as quantified by mean pre-pandemic Gross Domestic Product from 2015-2019 weighted by Purchasing Power Parity per capita (x-axis). Each dot represents a country-year datapoint, e.g., Senegal in 2021. For Panels A and B, dot colour indicates year of the pandemic – red for 2020, green for 2021, and yellow for 2022. Panel A (Per year) shows three lines of best fit for each year of the pandemic; Panel B (Combined) fits a single line of best fit for all years of data; and Panel C (Cumulative) plots total coverage deltas (the sum of each year). All fitted lines in the figure represent Loess (locally estimated scatterplot smoothing) curves, with the surrounding grey areas indicating the 95% confidence intervals for the fitted values. The horizontal black line indicates no change in DTP3 coverage. The vertical purple line indicates the visually-assessed threshold turning point.

D. **Stepwise linear regression results**

Linear regression models were built incrementally by adding explanatory variables according to their proximity to RI coverage as defined by our theoretical framework. Results from each step are detailed below in Table 1, alongside key statistics which informed whether we included the added variables in the next step for analysis.

**Table 1**: Stepwise linear regression modelling results

| **Step** | **Variables added/removed** | **BIC** | **Adjusted R^2^** | ***p*-value of new variables** | **Residuals** |
| --- | --- | --- | --- | --- | --- |
| 1 | Pre-pandemic immunisation programme strength | -1052.1 | 0.020 | 0.007 | Non-linear with turning point |
| 1a | **Added**: Threshold (>83% coverage)  **Added**: Interaction term | -1066.1 | 0.089 | < 0.00001  < 0.00001 | N/A |
| 1b | **Removed**: Pre-pandemic immunisation programme strength | -1071.9 | 0.089 |  | N/A |
| 2 | Health workforce | -1090.3 | 0.141 | < 0.00001 | Non-linear with turning point |
| 2a | **Added**: Threshold (> 60 health workers / 10k population)  **Added**: Interaction term | -1102.8 | 0.193 | 0.81  0.0004 | N/A |
| 2b | **Removed**: Health workforce threshold | -1108.6 | 0.196 |  | N/A |
| 3 | Health systems strength | -1103.0 | 0.194 | 0.060 | Linear |
| 4 | Global health security | -1102.8 | 0.193 | 0.086 | Linear |
| 5 | COVID-19 vaccination | -1102.9 | 0.194 | 0.74 | Linear |
| 6 | COVID-19 burden | -1108.0 | 0.21 | 0.02 | Linear |
| 7 | Health policy PC1  Health policy PC2 | -1098.9 | 0.195 | 0.16  0.94 | Linear  Linear |
| 8 | Government health expenditure | -1103.7 | 0.195 | 0.33 | Linear |
| 9 | External health expenditure | -1106.3 | 0.201 | 0.063 | Linear |
| 10 | Private health expenditure | -1103.1 | 0.194 | 0.54 | Linear |
| 11 | Outside mobility | -1108.0 | 0.205 | 0.023 | Linear |
| 12 | Residential mobility | -1103.6 | 0.195 | 0.36 | Linear |
| 13 | Containment policy | -1103.8 | 0.196 | 0.31 | Linear |
| 14 | Economic policy | -1103.0 | 0.194 | 0.62 | Linear |
| 15 | Country wealth | -1105.2 | 0.199 | 0.12 | Linear |

E. **Further Discussion**

High internal correlation between some predictors, as evidenced by Variance Inflation Factor (VIF) diagnostic analysis, may explain why Random Forest identified additional important variables compared to the linear regression, since collinearity can complicate interpretation of individual effects. VIF scores of over 5 indicate high correlation and this applied to: health workforce capacity (VIF = 5.61), government health financing (6.84), and health system strength (5.00). Country wealth also exhibited relatively high VIF score (4.69).

F. **Reproducibility**

All datasets are publicly available in our Git Hub repository [29] alongside the scripts for all analyses, which were conducted in R [30]. Results can be reproduced through running the scripts using the simplified ‘*reportfactory’* [31] commands. Key packages include: ‘*tidyverse*’ and ‘*ggplot2*’ for data cleaning and visualisation [32], ‘*stats*’ for PCA and linear regression [30], ‘*caret*’ for Random Forest [33].

[6]	World Health Organisation. The World Health Report 2006: Working Together for Health. 2006.

[7]	World Health Organization. The National Health Workforce Accounts database n.d. https://www.who.int/data/gho/data/themes/topics/health-workforce (accessed August 7, 2024).

[8]	World Health Organisation. Monitoring the building blocks of health systems: a handbook of indicators and their measurement strategies. 2010.

[9]	World Health Organisation. Coverage of essential health services (index based on tracer interventions that include reproductive, maternal, newborn and child health, infectious diseases, non-communicable diseases and service capacity and access) (SDG 3.8.1) n.d. https://platform.who.int/data/maternal-newborn-child-adolescent-ageing/indicator-explorer-new/MCA/coverage-of-essential-health-services-(index-interventions-reproductive-maternal-newborn-and-child-health)-(sdg-3.8.1) (accessed March 5, 2024).

[10]	World Health Organisation. Health security n.d. https://www.who.int/health-topics/health-security#tab=tab_1 (accessed March 5, 2024).

[11]	Global Health Security Index. GHS Index n.d. https://ghsindex.org/about/ (accessed February 28, 2024).

[12]	Economist Impact. GHS Index Methodology. n.d.

[13]	World Health Organisation. Third round of the global pulse survey on continuity of essential health services during the COVID-19 pandemic. 2022.

[14]	United Nations. World Population Prospects - Population Division - United Nations 2024. https://population.un.org/wpp/ (accessed August 25, 2024).

[15]	World Health Organisation. Pulse survey on continuity of essential health services during the COVID-19 pandemic: interim report, 27 August 2020. 2020.

[16]	World Health Organisation. Second round of the national pulse survey on continuity of essential health services during the COVID-19 pandemic. 2021.

[17]	Pinto AMC, Shariq S, Ranasinghe L, Budhathoki SS, Skirrow H, Whittaker E, et al. Reasons for reductions in routine childhood immunisation uptake during the COVID-19 pandemic in low- and middle-income countries: A systematic review. PLOS Global Public Health 2023;3:e0001415. https://doi.org/10.1371/JOURNAL.PGPH.0001415.

[18]	Karlinsky A, Kobak D. Tracking excess mortality across countries during the COVID-19 pandemic with the World Mortality Dataset. Elife 2021;10. https://doi.org/10.7554/ELIFE.69336.

[19]	World Health Organisation. Methods for estimating the excess mortality associated with the COVID-19 pandemic. n.d.

[30]	R Core Team. R: A Language and Environment for Statistical Computing. Vienna, Austria: 2021.

[31]	Jombart T, Gimma A, Taylor T. Lightweight Infrastructure for Handling Multiple R Markdown Documents [R package reportfactory version 0.4.0] 2021. https://doi.org/10.32614/CRAN.PACKAGE.REPORTFACTORY.

[32]	Wickham H, Averick M, Bryan J, Chang W, D’ L, Mcgowan A, et al. Welcome to the Tidyverse. J Open Source Softw 2019;4:1686. https://doi.org/10.21105/JOSS.01686.

[33]	Kuhn M. Building Predictive Models in R Using the caret Package. J Stat Softw 2008;28:1–26. https://doi.org/10.18637/JSS.V028.I05.
