## Supplementary material for "Pre-pandemic national immunisation programme strength and health workforce capacity improved routine immunisation resilience during the COVID-19 pandemic": Guideline checklist

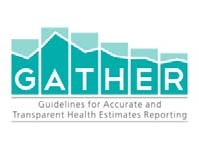
**Checklist of information that should be included in new reports of global health estimates**

| **Item #** | **Checklist item** | **Reported on page #** |
| --- | --- | --- |
| **Objectives and funding** | |  |
| **1** | Define the indicator(s), populations (including age, sex, and geographic entities), and time period(s) for which estimates were made. | Table 1 (section 2.2.2, pgs. 6-10) and Supplementary Materials Section B |
| **2** | List the funding sources for the work. | Acknowledgements (page 20) |
| **Data Inputs** | |  |
| *For all data inputs from multiple sources that are synthesized as part of the study:* | |  |
| **3** | Describe how the data were identified and how the data were accessed. | Table 1 (section 2.2.2, pgs. 6-10) and Supplementary Materials Section B |
| **4** | Specify the inclusion and exclusion criteria. Identify all ad‐hoc exclusions. | Table 1 (section 2.2.2, pgs. 6-10) and Supplementary Materials Section B |
| **5** | Provide information on all included data sources and their main characteristics. For each data source used, report reference information or contact name/institution, population represented, data collection method, year(s) of data collection, sex and age range, diagnostic criteria or measurement method, and sample size, as relevant. | Table 1 (section 2.2.2, pgs. 6-10) and Supplementary Materials Section B |
| **6** | Identify and describe any categories of input data that have potentially important biases (e.g., based on characteristics listed in item 5). | Table 1 (section 2.2.2, pgs. 6-10) and Supplementary Materials Section B |
| *For data inputs that contribute to the analysis but were not synthesized as part of the study:* | |  |
| **7** | Describe and give sources for any other data inputs. | Section 2.2.1 (pg.5) and Table 1 (section 2.2.2, pgs. 6-10) and Supplementary Materials Section B |
| *For all data inputs:* | |  |
| **8** | Provide all data inputs in a file format from which data can be efficiently extracted (e.g., a spreadsheet rather than a PDF), including all relevant meta‐data listed in item 5. For any data inputs that cannot be shared because of ethical or legal reasons, such as third‐party ownership, provide a contact name or the name of the institution that retains the right to the data. | All publicly available data, available on our [GitHub repository](https://github.com/bevans249/pandemic_RI_coverage_determinants) |
| **Data analysis** | |  |
| **9** | Provide a conceptual overview of the data analysis method. A diagram may be helpful. | Figure 1 (pg. 5) |
| **10** | Provide a detailed description of all steps of the analysis, including mathematical formulae. This description should cover, as relevant, data cleaning, data pre‐processing, data adjustments and weighting of data sources, and mathematical or statistical model(s). | Sections 2.2.3, 2.3, 2.4, and 2.5 (pgs. 11-13) |
| **11** | Describe how candidate models were evaluated and how the final model(s) were selected. | Section 2.4 (pgs. 10-11) and Supplementary Materials Section C and D |
| **12** | Provide the results of an evaluation of model performance, if done, as well as the results of any relevant sensitivity analysis. | Section 3 (pgs. 12-14) and Supplementary Materials Section C and D |
| **13** | Describe methods for calculating uncertainty of the estimates. State which sources of uncertainty were, and were not, accounted for in the uncertainty analysis. | Section 3 (pgs. 12-14) |
| **14** | State how analytic or statistical source code used to generate estimates can be accessed. | All publicly available data and scripts, available on our [GitHub repository](https://github.com/bevans249/pandemic_RI_coverage_determinants) |
| **Results and Discussion** | |  |
| **15** | Provide published estimates in a file format from which data can be efficiently extracted. | All publicly available data and scripts, available on our [GitHub repository](https://github.com/bevans249/pandemic_RI_coverage_determinants) |
| **16** | Report a quantitative measure of the uncertainty of the estimates (e.g. uncertainty intervals). | Section 3 (pgs. 12-14) and Supplementary Materials Section D |
| **17** | Interpret results in light of existing evidence. If updating a previous set of estimates, describe the reasons for changes in estimates. | Section 5 (pgs. 15-19) |
| **18** | Discuss limitations of the estimates. Include a discussion of any modelling assumptions or data limitations that affect interpretation of the estimates. | Section 5 (pgs. 18-19) |

*This checklist should be used in conjunction with the GATHER statement and Explanation and Elaboration document, found on gather‐statement.org*

1
